# Personalized Definition of Short Sleep Using Long-Term Wearable Sleep Distributions

**DOI:** 10.64898/2026.07.06.26357372

**Authors:** Chun Siong Soon, Xin Yu Chua, Shuo Qin, Ju Lynn Ong, Stijn A. A. Massar, Adrian Willoughby, Kyra H. M. Chong, Michael W. L. Chee

**Affiliations:** Centre for Sleep and Cognition, Yong Loo Lin School of Medicine, National University of Singapore (NUS), Singapore; Oura-National University of Singapore (NUS) Joint Lab, Oura Pte Ltd, Singapore

**Keywords:** Sleep duration, short sleep, inadequate sleep, time series modeling, sleep variability, cardiometabolic health, ecological momentary assessment, wearable devices, data-driven personalization, sleep homeostasis

## Abstract

**Study Objectives:** To evaluate a framework using wearable data to personalize the definition of ‘short sleep’, comparing its temporal and functional characteristics against a fixed threshold.

**Methods:** 462 healthy adults wore sleep trackers and provided daily ecological momentary assessments for a year. Short sleep was defined using either a fixed threshold of <6 h/night (fSS) or personalized thresholds anchored to individual sleep-duration distributions (pSS). Temporal patterns of consecutive short-sleep nights were characterized. Linear mixed-effects models examined associations between accumulating short-sleep nights and short- and long-term markers. Sleep patterns across six other countries were also evaluated.

**Results:** pSS and fSS produced similar average thresholds and overall prevalence of short-sleep nights. However, pSS showed larger effect estimates for short-term outcomes, including alertness, sleep satisfaction, stress, sleep heart rate, HRV, and sedentariness. Effects increased with successive short-sleep nights. Proportion of pSS showed stronger association with blood pressure and arterial stiffness. Isolated short nights were common, whereas longer runs were uncommon and typically followed by incomplete recovery sleep. Personalized thresholds distinguished stable short sleepers with few pSS nights from individuals experiencing recurrent sleep shortfall and highlighted vulnerability among those achieving ‘recommended’ sleep duration but with high variability. Despite marked cross-country differences in sleep habits, the distribution of short-sleep runs, and termination patterns were remarkably similar.

**Conclusion:** Anchoring short sleep to an individual’s habitual sleep distribution captures relative sleep shortfall beyond absolute duration, better characterizing the functional impact of ‘short’ sleep. Preventive strategies may benefit from limiting pSS accumulation together with addressing sporadic inadequate sleep.

**Statement of Significance:** Evidence describing how short sleep occurs under real-world conditions remains limited. Using long-term wearable recordings, we show that isolated short nights are common whereas extended runs are uncommon and typically followed by recovery sleep. We evaluated a personalized definition of inadequate sleep anchored to individual habitual sleep patterns rather than a fixed duration threshold. This framework more sensitively captured short-term functional impairments involving alertness, stress, sleep satisfaction, autonomic function and sedentariness. Stronger associations with longer-term vascular risk markers were also observed. Personalized wearable-derived metrics may provide a scalable approach to individualizing sleep planning without undue emphasis on occasional short sleep.

## Introduction

Adequate and regular sleep is a core determinant of cognitive, emotional and physical health^1,2^. Current sleep duration guidelines provide age-specific, range-based recommendations^3^ that are heavily weighted by self-reported data^4^. In practice, many working adults aim to remain above the lower bound of these recommendations to avoid short sleep (SS), balancing daytime alertness, performance, mood, and physiological functioning against competing demands on waking time. The increasing use of consumer health trackers has shifted how sleep is measured and interpreted^5,6^, raising questions about how population-based recommendations apply personally to objectively estimated sleep in everyday settings.

Across multiple studies, short sleep is most commonly operationalized using a fixed threshold of less than six hours per night^7–9^. While this fixed short sleep (fSS) definition has utility for population-level messaging, it has several limitations in the context of continuous digital measurement. First, fSS is largely based on self-report, whereas PSG or wearable derived measurement typically yield shorter sleep durations^10,11^. Sleep health guidelines may acknowledge this discrepancy but do not provide a practical solution. Second, fixed thresholds do not account for inter-individual variation in habitual sleep duration^12^. Third, fixed definitions treat short sleep as an isolated exposure and do not capture its temporal structure in real life, including the frequency of occurrence, clustering into consecutive runs, and constraints on recovery sleep.

Experimental sleep restriction studies^13–15^ and a large observational study^16^ show that the functional consequences of shortened sleep accumulate across successive nights and that more days of recovery sleep may be required to resolve incurred deficits^15,17–20^. However, the long-term pattern of short sleep under free-living conditions—beyond weekday-weekend differences and social jetlag^21,22^ —has rarely been characterized at scale using objective data^23^. In particular, it remains unclear within individuals how often short sleep occurs, the characteristics of short-sleep runs, and how these patterns relate to short-term functional outcomes and longer-term cardiometabolic risk.

Longitudinal sleep data from healthy individuals using wearable health trackers provide an opportunity to address these gaps. Such data enable characterization of SS as it naturally occurs^23^ and evaluation of how deviations from habitual sleep relate to next-day wellbeing^24^, cardiac autonomic markers^25^, sleep satisfaction and sedentariness^26^.

The primary aims of this study were to characterize both within and between individuals, the frequency and temporal structure of short sleep in a large cohort of working adults, and to examine the length and functional consequences of consecutive short-sleep sequences.

We evaluated a personalized definition of short sleep (pSS) derived from individual sleep-duration distributions. We reasoned that a personalized definition of short sleep should reflect a functionally meaningful shortfall relative to an individual’s habitual sleep pattern rather than merely below-average sleep. Accordingly, we evaluated a personalized framework anchored to the upper portion of individual sleep-duration distributions and incorporating an absolute decrement informed by experimental sleep restriction literature. We hypothesized that this approach would provide greater functional sensitivity than a fixed threshold for detecting the effects of accumulating short sleep on day-to-day wellbeing and cardiovascular markers. Finally, we examined whether fixed and personalized definitions of short sleep differed in their associations with longer-term markers of obesity and vascular health.

## Methods

### Participants and design

In this year-long observational study, mostly healthy full-time staff members from the National University of Singapore (NUS) were recruited between October 2024 and January 2025 via university-wide email announcements, posters, and word-of-mouth referrals. After online screening, eligible participants (age 21–70 years; not engaged in shift work; not pregnant or breastfeeding; no self-reported history of sleep, psychological, or neurological disorders) provided written informed consent. The study protocol was approved by the NUS Institutional Review Board and complied with the Declaration of Helsinki. Participants received up to SGD182 (∼USD 140), prorated by completion rate. Those who completed ≥80% of daily measures could retain the Oura Ring.

### Study procedures

Baseline cardiometabolic assessments were performed after consent was provided to assess cardiometabolic health. Participants then contributed daily Oura Ring (Gen 3; Oura Health Oy, Oulu, Finland) and smartphone-based ecological momentary assessment (EMA) data for up to 12 months. Only participants who provided at least 28 days of concurrent Oura Ring and EMA data were included in analyses. Study measures not relevant to the present analyses are omitted. Unless otherwise stated, descriptive statistics are reported as median [interquartile range (IQR)].

#### Baseline cardiometabolic assessments

To assess cardiovascular health, measurements of aortic or central pulse pressure (CPP, the difference between central systolic and diastolic blood pressure), carotid-femoral pulse wave velocity (PWV), and augmentation index (Alx, the portion of the pressure wave from the heart reflected back from small arteries) were made using a tonometer (SphygmoCor XCEL, Cardiex Ltd). In addition, an arterial stiffness composite score was calculated by averaging the cohort-normalized z-scores of CPP, PWV and Alx^27^. All other cardiometabolic outcomes were analyzed in their original units.

Obesity was assessed using the body roundness index (BRI)^28^:

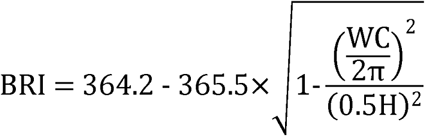

where WC denotes waist circumference (cm) and H denotes height (cm).

#### Wearable Device Usage

Participants were instructed to wear an Oura Ring Gen 3 (Oura Health Oy, Oulu, Finland) for a minimum of 20 hours per day to monitor their sleep and activity. Nightly mean heartrate (HR) and heartrate variability (HRV) during sleep were recorded. The accuracy of these measures was previously validated^29^.

For each day, the main sleep period was defined as the longest sleep period occurring between 18:00 on the previous day and 17:59 on the current day, while additional sleep periods within this window were classified as naps. To ensure valid characterization of sleep under free-living conditions, predefined exclusion criteria based on device wear-time and uncertainty regarding capture of the main sleep episode were applied. Periods of overseas travel, collated through self-reports and Oura-detected time zone changes, were excluded. This resulted in the exclusion of 7,144 nights (5.25%; See Supplementary Methods). Missing data were not imputed as there was an abundance of data.

#### Daily EMA

EMA data were collected daily using a mobile application (Z4IP, Sleep and Cognition Lab, Singapore). To encourage continued daily participation over the course of a year in working adults, the study protocols were designed to minimize participant burden. Participants completed an EMA once a day, between 19:00 and 23:45. They were prompted through smartphone notifications to complete a questionnaire with items on emotions (visual analogue scale from 1–*Not at all* to 100 – *Extremely* for “Alert/Attentive”, “Motivated”, “Sad / Down” and “Stressed”), sleep satisfaction referring to the previous night’s sleep (Likert scale from 1– Very poorly to 5 – Very well), and caffeine intake (select all 4-hour time windows from prior day 20:00 to current day 19:59 during which any caffeinated drink was consumed). Caffeine intake was operationalized as the number of 4-hour windows in which caffeinated drinks were consumed (range 0–6) and included as a covariate in mixed-effects models.

#### Sleep Regularity Index (SRI)

SRI was calculated based on Phillips et al. (2017)^30^ but using 15min instead of 30s windows:

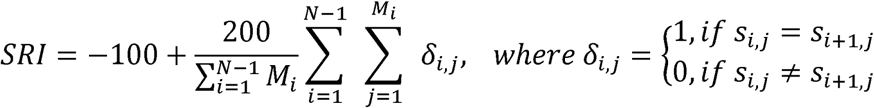

*s_i,j_* is the state of arousal for a 15min window *j* on day *i* SRI was calculated based on pairs of consecutive nights, and non-wear periods on either day were excluded, i.e., *M_i_* included only valid windows where neither *s_i,j_* nor *s_i+1,j_* was missing. SRI calculation included all sleep episodes in a day.

#### Short sleep run identification

Daily sleep duration was obtained from the Oura Ring and quantified using two metrics: (i) main sleep period duration (total sleep time; TST) and (ii) 24h TST (combined duration of all additional sleep episodes; see Supplementary Methods). Short sleep nights were identified using: (i) a fixed short-sleep threshold (fSS), defined as TST <6.0 h, commonly used in sleep literature; and (ii) a personalized short-sleep threshold (pSS), defined as TST <P75 − 1.0 h, where P75 denotes the 75th percentile of the participant’s TST across the study period, to account for inter-individual differences in habitual sleep duration. More stringent thresholds (“Very Short Sleep”; Table S5) were also examined: (i) fixed, 5.5 h; and (ii) personalized, P75 − 1.5 h.

The personalized framework combined an individualized percentile reference with an absolute decrement. A percentile criterion alone risks classifying a fixed proportion of nights as short irrespective of functional significance, whereas purely variance-based approaches may yield thresholds that are excessively permissive or overly stringent depending on sleep regularity. Higher percentiles of an individual’s sleep-duration distribution preferentially reflect nights with fewer occupational and social constraints and therefore provide a practical approximation of realized higher-opportunity sleep. Subtracting one hour from this reference was intended to represent a functionally meaningful sleep shortfall, informed by experimental sleep restriction findings demonstrating measurable effects of approximately one hour of restriction on vigilance, mood, and wellbeing.

Because participants contributed differing numbers of recording days, opportunity for short sleep could vary across individuals. Propensity for short-sleep could also affect participation duration. To evaluate these possibilities, Spearman’s correlations were computed between number of recorded days and proportion of short-sleep nights.

For each short-sleep threshold, runs of consecutive short-sleep nights were identified (minimum run length = 1). To ensure a valid non-short-sleep baseline, a short-sleep run was required to be preceded by a non-short-sleep night. Because it cannot be determined whether a missing night qualified as short sleep, runs immediately following missing nights were excluded from analysis. Numbers of excluded runs are reported in the Supplementary Results. These filtering procedures remove ambiguity concerning sequence history that could otherwise confound interpretation and are feasible in long-duration wearable datasets comprising hundreds of nights while preserving inferential power.

#### State effects: Multi-domain associations of consecutive nights of short sleep

Linear mixed-effects models tested whether the number of consecutive nights of short sleep experienced, i.e., the day index within a short sleep sequence, was associated with day-to-day variations in self-reported wellbeing and physiological markers (fitlme function in MATLAB R2024a; MathWorks, Natick, MA, USA):

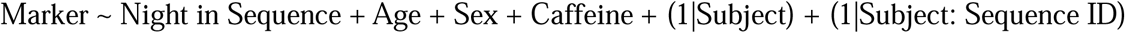

Days not classified as short sleep were assigned an index of 0. Age, sex, and caffeine intake were included as covariates. Random intercepts for participants accounted for repeated measures and individual baseline differences, while an additional random intercept for sequence ID nested within participant accounted for the non-independence of observations within the same short-sleep sequence.

Both unstandardized and standardized beta coefficients were reported. Unstandardized coefficients were included to facilitate interpretation in the original measurement units, while standardized coefficients were additionally reported to aid comparison of the relative strength of associations across outcomes and SS threshold definitions (fSS vs. pSS)

Outcome markers included daily motivation, sadness, stress, alertness, sleep satisfaction, physical movement, sedentary time, as well as sleeping HR and HRV.

#### Trait effects: Associations with cardiometabolic health

Linear regression models evaluated the trait effects of each participant’s proportion of short sleep, as defined by fSS and pSS, on cardiometabolic health markers, including BRI, PWV, systolic blood pressure and arterial stiffness composite score (MATLAB R2024a; MathWorks, Natick, Massachusetts):

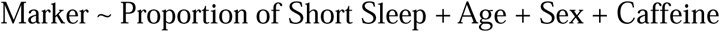

Both unstandardized and standardized beta coefficients were reported. Unstandardized coefficients were included to facilitate interpretation in the original measurement units, while standardized coefficients were additionally reported to aid comparison of the relative strength of associations across outcomes and SS threshold definitions (fSS vs. pSS)

#### Recovery sleep analyses

We examined the probability of short sleep on weekdays versus weekends, and each day of the week. In addition, we quantified recovery sleep duration as the duration on the first night following the last night of a short sleep run. This would be the night following the first and only night of runs of length one and following the 5th night of runs of length 5. Separate analyses were conducted depending on whether the recovery night occurred on a weekday or on a weekend.

#### Multi-country supporting data

The multi-country supporting data to support the generalizability of the proposed sleep framework was provided by Ouraring Inc (San Francisco, CA) according to their terms and conditions (https://ouraring.com/terms-and-conditions), and privacy policy (https://ouraring.com/en/privacy-policy-oura-health). Data used for this study were anonymized or aggregated consistent with Oura’s privacy policy. The data come from users in 7 countries with different sleep habits (Australia, Finland, Germany, Japan, Singapore (separate from the data reported here with a broader sample), the United Kingdom and the United States of America). Data from 430,386 users were collected from 1st March 2024 to 28th February 2025. Mean age was 42.6 years (SD=13.2), and the proportion of females was 66.2% (see Table S11). Users were required to have at least 100 nights of valid sleep recordings to be included. The definition of fSS and pSS as well as the recovery sleep analysis followed the procedure described for the main study. This data does not have functional marker information.

## Results

### Participant characteristics and sample-average sleep parameters

575 participants (median [IQR] age: 39.8 [16.2] years; 351 [61.0%] females) enrolled in this study. Of these, 462 participants (median [IQR] age: 40.4 [15.9] years; 283 [61.3%] females) with at least 28 days of concurrent Oura Ring and EMA data were included in the analyses (median [IQR] 212.5 [172.0] days).

Across the sample, sleep patterns were on average in steady state over the year (Fig. S1), showing a range of 30-40 minutes dominated by workday versus free day (weekends and public holidays) sleep duration differences.

Within-person median TST was (median [IQR]) 6.29 [0.95] h, showing a quasi-normal distribution (Table 1; Fig. 1a). Within-person sleep variability measured by standard deviation of TST was (median [IQR]) 1.08 [0.39] h. Sleep regularity assessed by SRI was 76.53 [8.55]. Individual napping rates were higher (*t*_461_=20.9, *p*<.05) on weekends (median [IQR]: 37.4 [28.53] %) than on weekdays (19.53 [15.67] %). When they occurred, median weekend nap duration (44.0 [36.8] min) was also longer (*t*_461_=12.6, *p*<.05) than weekdays (29.5 [23.0] min). Nap distribution properties can be found in Supplementary Results (Fig. S3). Additional analyses found that naps did not affect the associations between consecutive nights of short sleep and functional markers and are not further discussed (see Table S3).

**Fig. 1.**
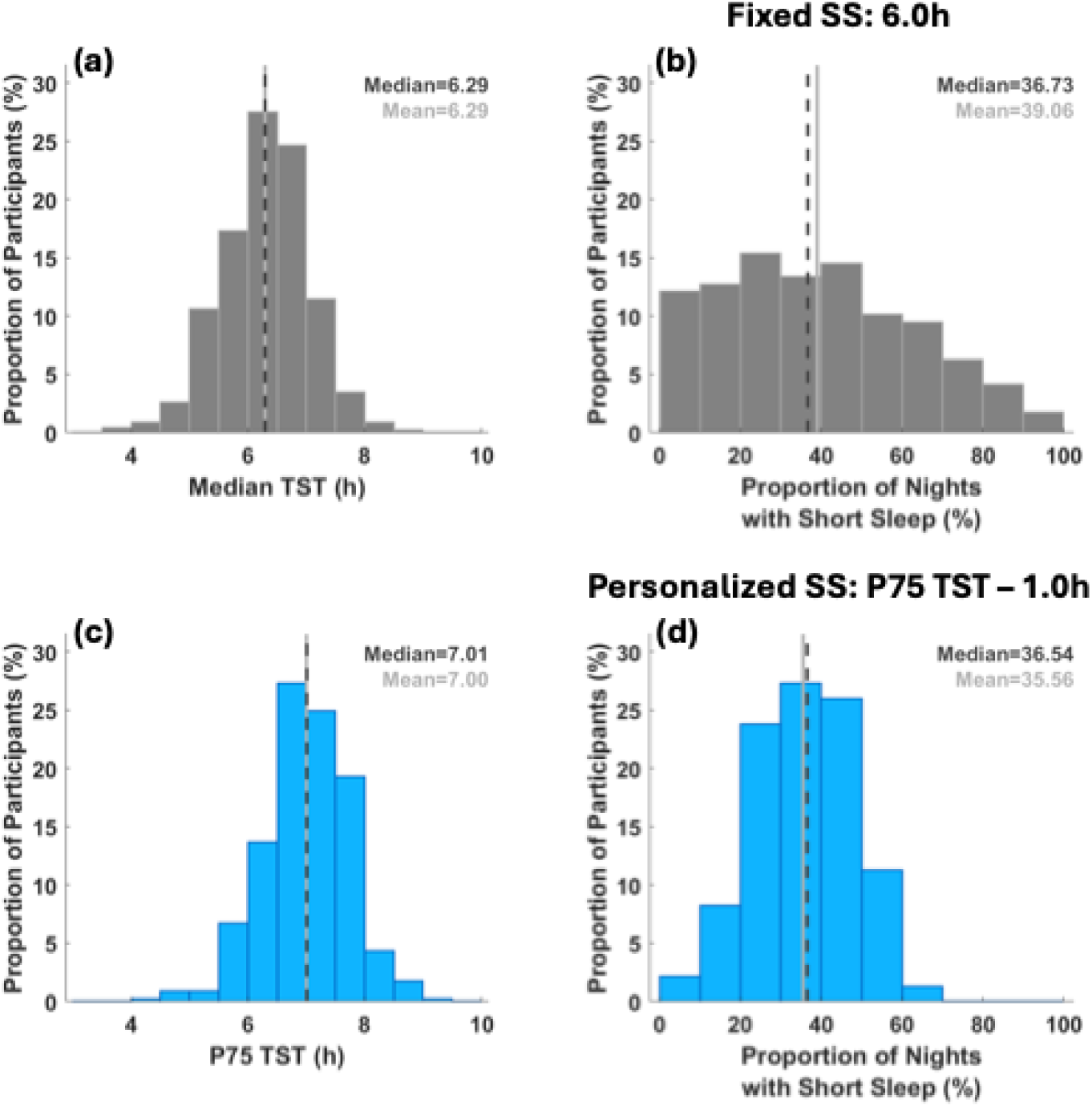
Distributional properties of fixed (6.0h) vs personalized (P75 − 1.0h) short sleep. (a) The median sleep duration of this cohort was relatively short at 6.4h (median), matching previous reports of sleep in Singapore. Notably, 27.6% of participants slept on average less than the fixed SS threshold of 6.0h. (c) The 75th percentile of each individual’s TST distribution was 7.1h (median), yielding a pSS threshold centered around 6.1h, close to the 6.0h fSS threshold. (b) vs (d): Comparable proportions of nights were classified as short sleep for fSS (median=35.3%) and pSS (median=36.9%), despite differences in the shape of the fSS and pSS distributions.

**Table 1.**
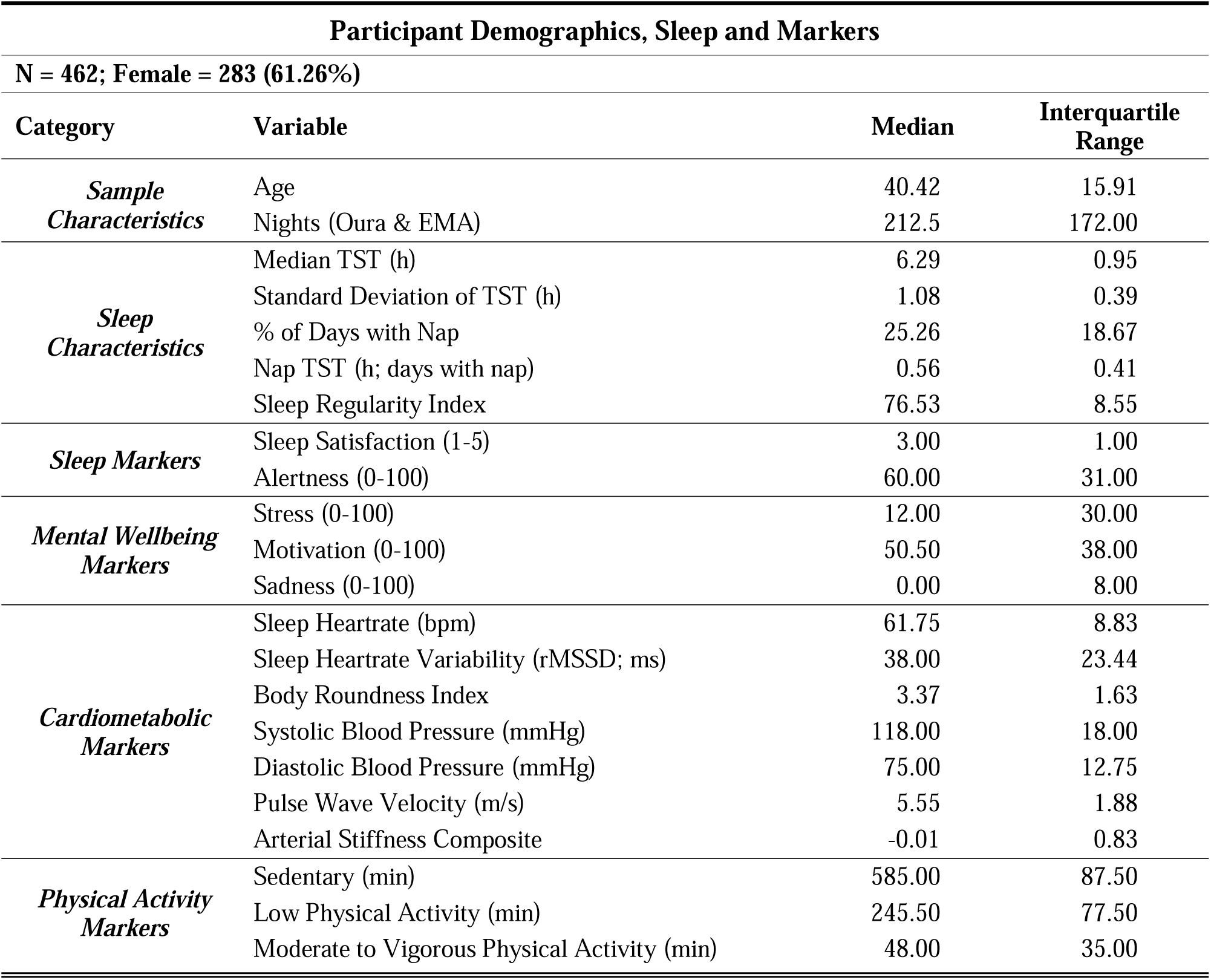
Participant Demographics, Sleep and Wellbeing Markers.

### Frequency of occurrence and sequential information about nights of ‘short sleep’

Defining SS as TST <6.0h per night, the average proportion of SS nights was 35.3% [34.1%]. 76.1% of individuals had a median run length of only 1 night. The distributions of proportion of SS nights and SS run length were heavily right-skewed (Fig. 1b; 2a-c), exposing large between-person differences: some people almost never have SS while others encounter it most of the time, consistent with the trait-like nature of sleep duration.

**Fig. 2.**
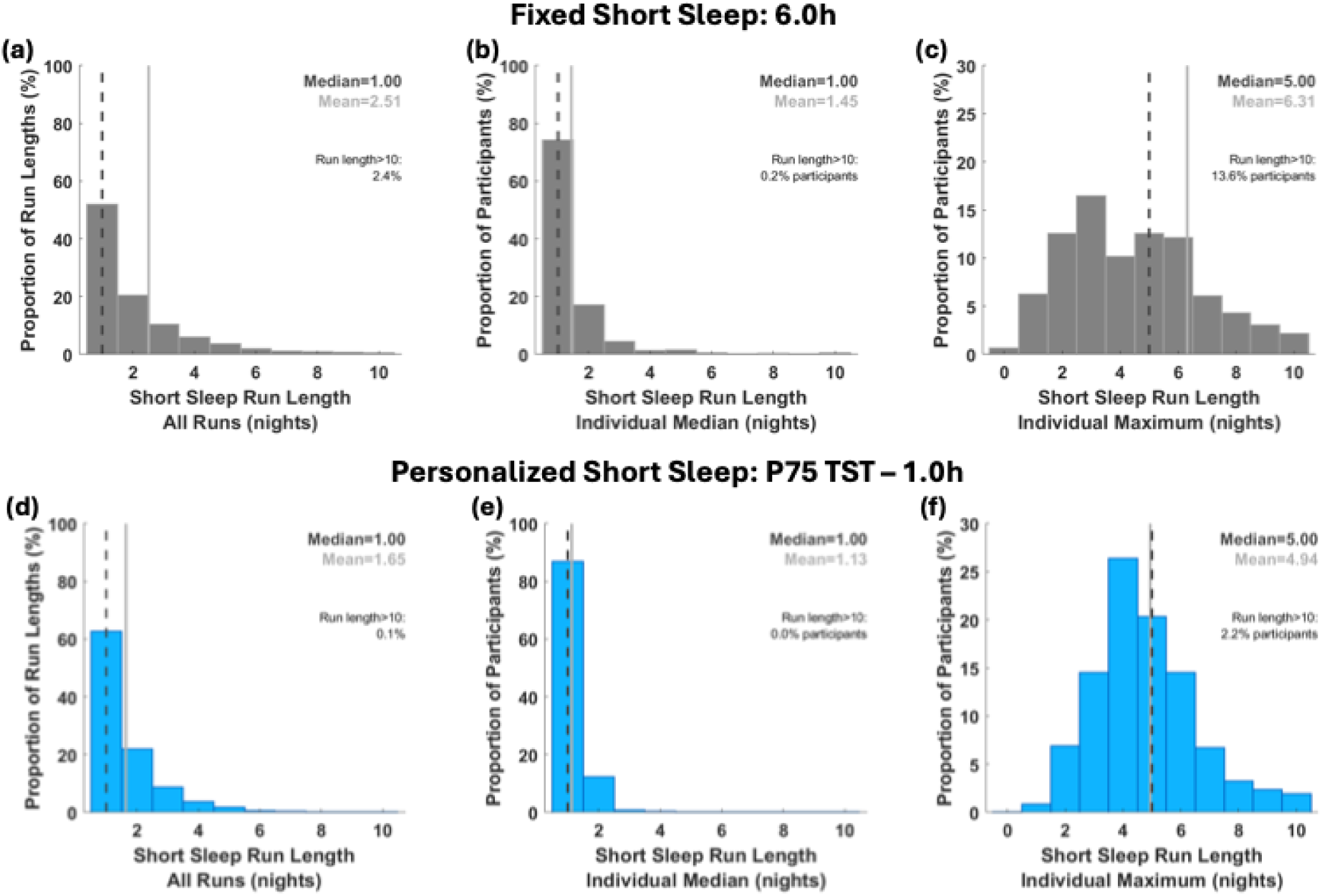
Short sleep run lengths (number of consecutive nights classified as SS) based on fSS and pSS thresholds. (a) and (d): For both fSS and pSS, the median run length across the cohort was one night, i.e., most SS runs lasted only one night. (b) and (e): Similarly, most individuals had a median run length of only one night for both fSS (76.1%) and pSS (86.4%). (c) and (f): Despite the short median run length, most participants experienced at least one occurrence of a run of consecutive SS nights. The median maximum run length for individuals was 4 nights for both fSS (mean=5.87) and pSS (mean=4.60).

The personalized threshold used was 1.0h less than the 75th percentile of an individual’s long run (at least a month) sleep duration (Fig. 2d-f). Using this personalized threshold did not affect either the group-level average duration of SS, 6.1 [0.7] h (median [IQR]), or the average proportion of nights classified as short: 36.9% [18.0%] (median [IQR]).

At the individual level, however, classification altered how shortfall in sleep was classified. Overall, 86.4% of individuals had a median run length of only 1 night (10.3% more than with the fSS definition). When pSS threshold was <6.0 h, fewer nights were classified as short compared to if fSS were used (fSS only: 21.7 (19.1)%; overlapping: 34.5 (11.8)%; Fig.4), This reclassifies individuals with a lower habitual sleep duration, for whom shorter sleep may be adequate. An extreme example of this is an individual with an average TST of <5h but a low SV of 0.5h and high SRI of 91.9 (Fig. 5). Conversely, persons sleeping habitually longer could be assessed to have more SS nights even though they have relatively more nights of >6h sleep (pSS only: 14.9 (9.6)%; overlapping: 22.1 (13.7)%), identifying nights of inadequate sleep among habitually longer sleepers (Fig. S2).

**Fig. 3.**
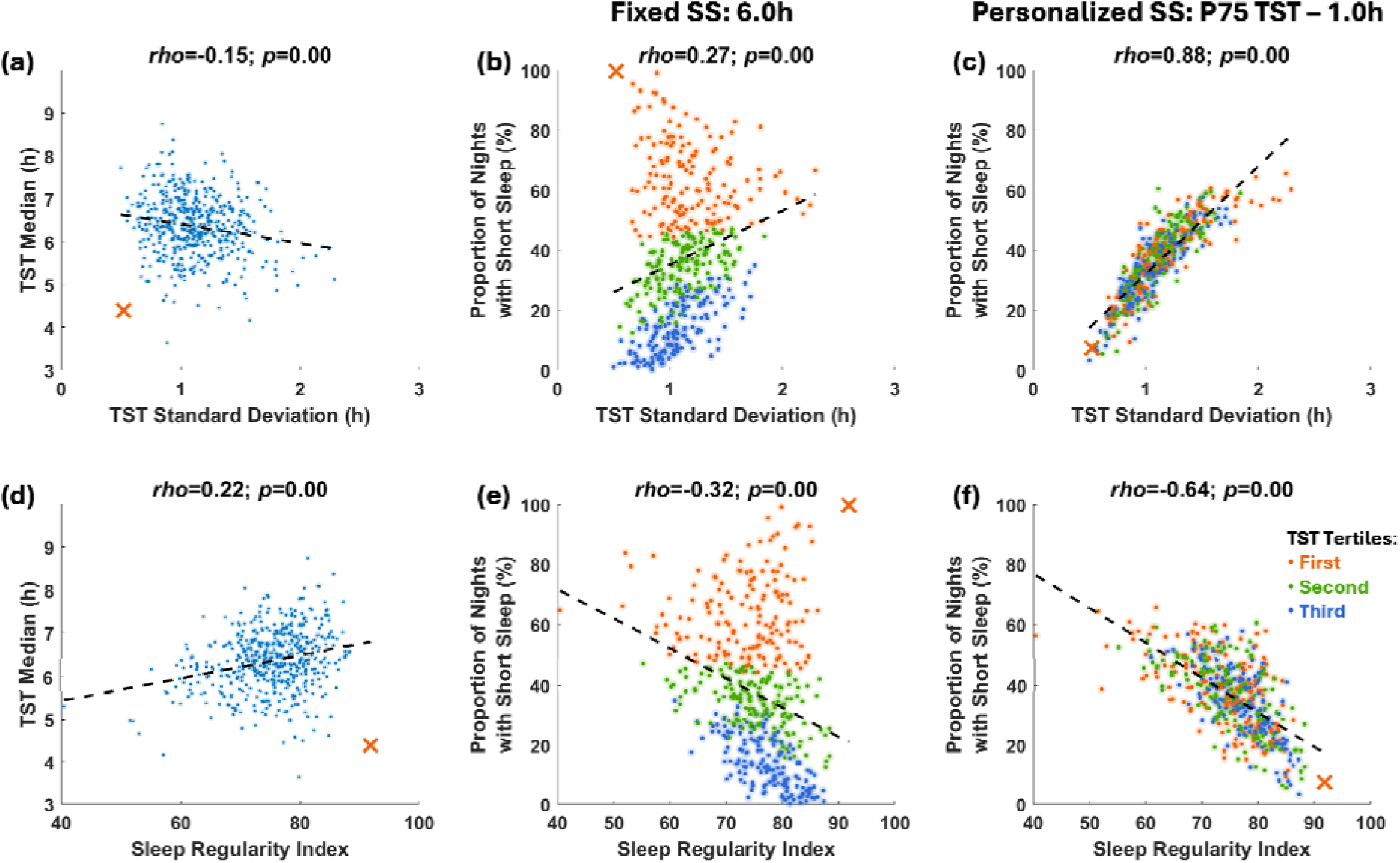
Associations between sleep duration, variability and regularity for different SS definitions. (a) and (d): Correlation of median sleep duration with sleep variability and regularity, (b), (c), (e) and (f): Colors code for membership within each tertile of median TST. Across all panels, the red **×** represents a short sleeper with low sleep variability and high regularity. In contrast to the high proportion of short sleep nights using fSS: 99.6%, the proportion of short sleep nights using the pSS definition was low, 7.4%; (see Fig. 4 for further details).

**Fig. 4.**
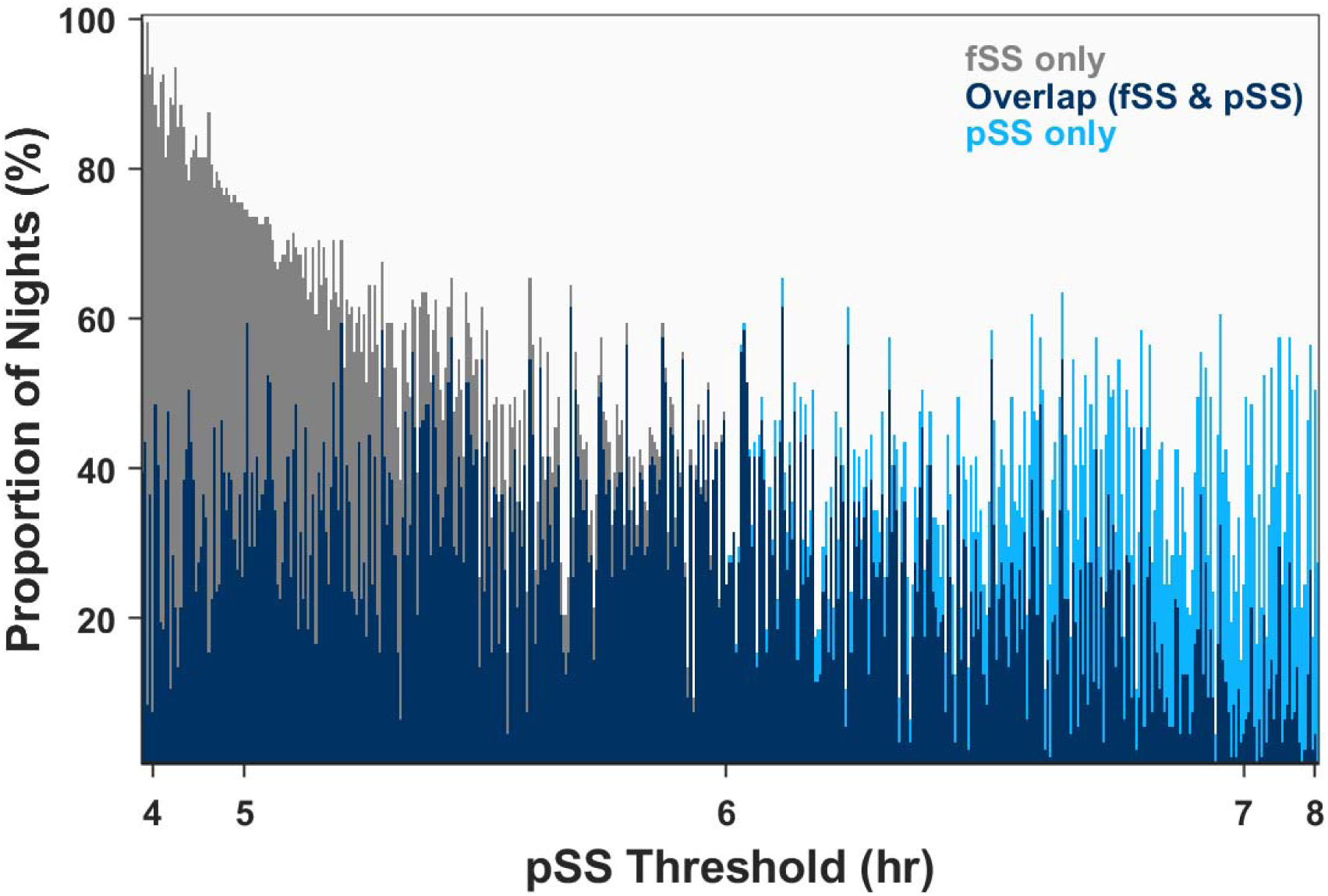
Proportion of nights classified as ‘short sleep’ according to a fixed <6h (fSS) threshold and a personalized (pSS) threshold. With the pSS threshold at <6.0 h, fewer nights were classified as short compared to if fSS were used (fSS only: 21.7 (19.1)%; overlapping: 34.5 (11.8)%), Conversely, persons sleeping habitually longer could be assessed to have more SS nights even though they have relatively more nights of >6h sleep (pSS only: 14.9 (9.6)%; overlapping: 22.1 (13.7)%), identifying nights of inadequate sleep among habitually longer sleepers.

**Fig. 5.**
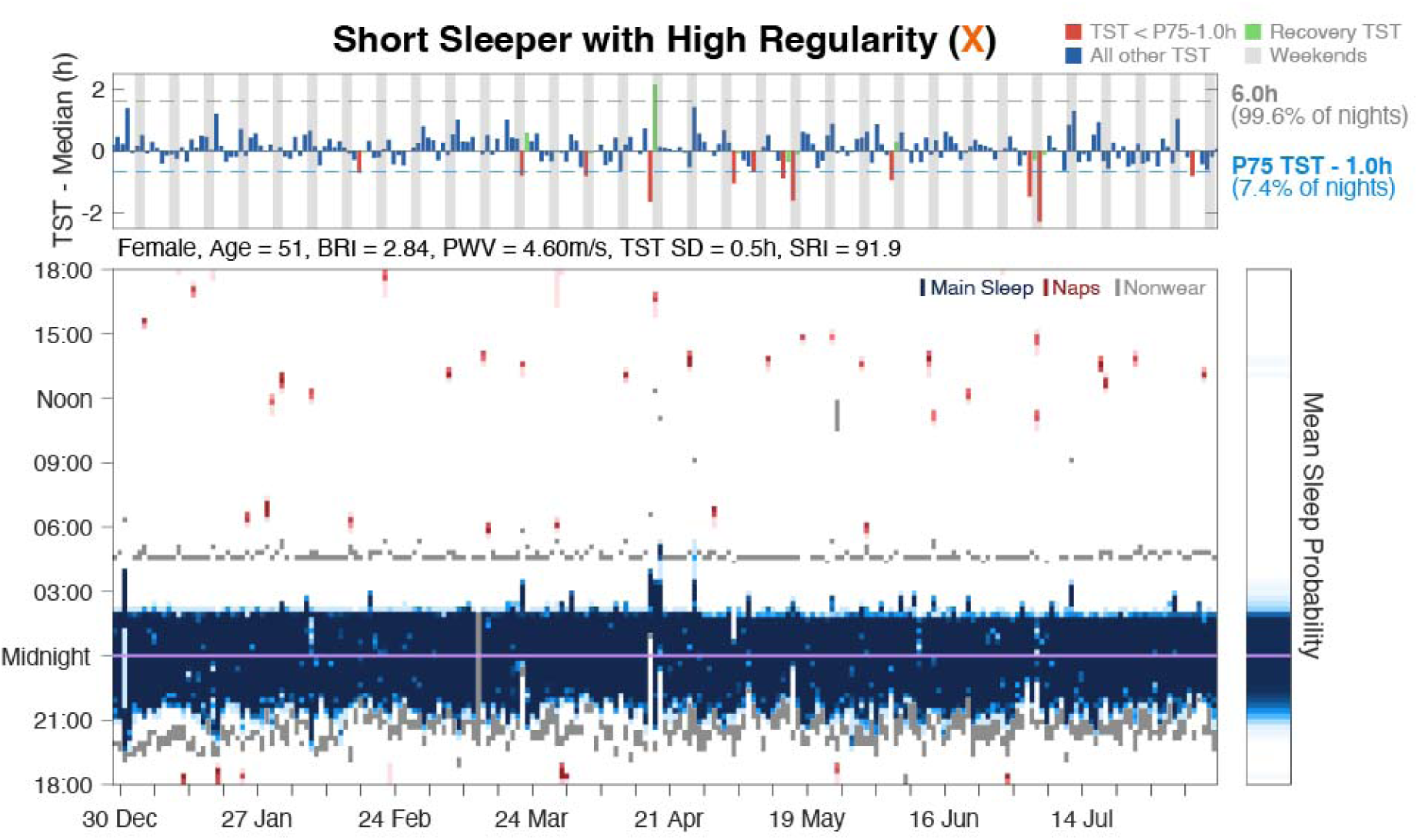
Sleep profile of an individual with many fSS nights but few pSS nights. This short sleeper (median TST=4.4h) with highly regular sleep patterns (SRI=91.9; TST IQR=0.7h) is marked by a red “×” in Fig. 3. Bottom panel shows daily main nocturnal sleep (blue), naps (red) and non-wear time (grey). Despite extremely short habitual sleep, this individual only napped occasionally (15.9%) for 19.0 [11.5] min (median [IQR]). Top panel shows daily main sleep duration relative to median TST. In contrast to a scarcity of nights (red) below the pSS threshold (blue dashed line), every night was below the fSS threshold (grey dashed line) except for one – a recovery night (green) following a night with just 2.1h of sleep. This person has a BRI of 2.84 and a PVW of 4.6 m/s which are healthy for a 51 year old.

The percentage of SS was not significantly correlated with the number of days of data for both fSS (Spearman’s *rho* = –.03, *p*=.49) and pSS (Spearman’s *rho* = –.07, *p*=.16), suggesting that participants with more days of data were not systematically more or less likely to experience short sleep, i.e. attrition rates were not biased by SS propensity.

### State effects: Next-day consequences of additional nights of short sleep

After accounting for age, sex, and caffeine intake as covariates, as well as participant-and sequence-level random intercepts, unstandardized regression coefficients were larger across several markers under the personalized short-sleep (pSS) definition compared with the fixed-threshold short-sleep (fSS) definition (Table 2). Standardized regression coefficients showed a similar pattern, indicating stronger relative associations under the personalized definition and suggesting that individualized short-sleep thresholds may better capture the magnitude of next-day functional alterations associated with accumulating short sleep.

**Table 2.** Personalized vs Fixed Definitions of Short Sleep: Associations Between Consecutive Days of Short Sleep.

| Marker ~ Night in Sequence* + Age + Sex + Caffeine + (1\|Subject) + (1\|Subject:Sequence ID) |  |  |  |  |  |
| --- | --- | --- | --- | --- | --- |
| Category | Marker | Fixed Short Sleep<br>(6.0h) |  | Personalized Short Sleep<br>(P75 TST – 1.0h) |  |
| | | *B*<br>[SE] | $\beta$<br>[SE] | *B*<br>[SE] | $\beta$<br>[SE] |
| *Mental Wellbeing Markers* | Stress<br>(Range: 0-100) | **0.196\*\*\***<br>[0.028] | **0.008\*\*\***<br>[0.001] | **0.794\*\*\***<br>[0.062] | **0.033\*\*\***<br>[0.003] |
|  | Motivation<br>(Range: 0-100) | **-0.334\*\*\***<br>[0.025] | **-0.012\*\*\***<br>[0.001] | **-0.608\*\*\***<br>[0.055] | **-0.022\*\*\***<br>[0.002] |
|  | Sadness<br>(Range: 0-100) | **0.065\*\***<br>[0.020] | **0.004\*\***<br>[0.001] | **0.223\*\*\***<br>[0.046] | **0.013\*\*\***<br>[0.003] |
| *Sleep Markers* | Sleep Satisfaction<br>(Range: 1-5) | **-0.022\*\*\***<br>[0.001] | **-0.023\*\*\***<br>[0.001] | **-0.118\*\*\***<br>[0.003] | **-0.123\*\*\***<br>[0.003] |
|  | Alertness<br>(Range: 0-100) | **-0.428\*\*\***<br>[0.022] | **-0.017\*\*\***<br>[0.001] | **-1.046\*\*\***<br>[0.048] | **-0.042\*\*\***<br>[0.002] |
| *Cardiovascular Markers* | Sleep Heartrate<br>(bpm) | **0.013\***<br>[0.006] | **0.002\***<br>[0.001] | **0.166\*\*\***<br>[0.015] | **0.020\*\*\***<br>[0.002] |
|  | Heartrate Variability<br>(rMSSD; ms) | **-0.037\***<br>[0.016] | **-0.002\***<br>[0.001] | **-0.288\*\*\***<br>[0.040] | **-0.013\*\*\***<br>[0.002] |
| *Physical Activity Markers* | Sedentary<br>(min) | **1.218\*\*\***<br>[0.177] | **0.010\*\*\***<br>[0.001] | **4.254\*\*\***<br>[0.425] | **0.033\*\*\***<br>[0.003] |
|  | Low Physical Activity<br>(min) | **0.999\*\*\***<br>[0.120] | **0.011\*\*\***<br>[0.001] | **4.896\*\*\***<br>[0.291] | **0.051\*\*\***<br>[0.003] |
|  | Moderate to Vigorous<br>Physical Activity (min) | **0.323\*\*\***<br>[0.060] | **0.007\*\*\***<br>[0.001] | **1.370\*\*\***<br>[0.144] | **0.029\*\*\***<br>[0.003] |

\* $p < .05$ ; \*\* $p < .005$ ; \*\*\* $p < .0005$
Sequence refers to a run of consecutive nights of ‘short’ sleep.

These findings remained consistent when the model was reanalyzed as a base model without covariates (Table S1) and after removing caffeine as a covariate (Table S2).

Adding SRI as a co-variate did not attenuate the day-in-sequence effect (Table S4), indicating that while proportion of nights below pSS strongly correlates with sleep variability (Fig. 2c), the effect of accumulating individualized short-sleep nights on next-day outcomes cannot be explained by sleep variability alone.

### Trait effects: Effect of short sleep on cardiometabolic markers

In the regression models, personalized short sleep (pSS) showed stronger associations with BRI, systolic blood pressure, pulse wave velocity and arterial stiffness composite than fixed short sleep (fSS), with findings remaining stable across sensitivity analyses using alternative personalized thresholds. The effect of age on these markers was notable (Fig. S5) but was accounted for.

Sensitivity analyses using alternative personalized thresholds (P80−1h and P75−1.5h) yielded congruent results, with pSS showing consistently larger associations with all markers than matched fixed-threshold definitions. This indicates that the relationship between pSS and vascular risk markers is robust to reasonable variation in threshold specification and is unlikely to reflect threshold tuning.

### Connection between short sleep defined by fixed vs. personalized approaches with sleep variability and regularity

In the multidimensional appraisal of sleep, duration and regularity are separate dimensions^31,32^. Sleep regularity (SR) and sleep variability (SV) are closely related but not merely the inverse of one another as the former as measured by SRI^33^ considers similarity in day-to-day sleep timing, duration and continuity while variability measured by the standard deviation of sleep duration measures only dispersion of sleep duration.

There was a weak correlation between average sleep duration and individual SV of ρ = −0.15, SRI of ρ = 0.22 (Spearman’s; Fig. 3a, d), suggesting that duration variability shares only limited variance with habitual duration. Using fSS yielded only a modest positive correlation (ρ = 0.27) for proportion of fSS nights and SV (Fig. 3b). This is explained by the heterogeneity of sleep duration at each tertile of frequency band of fSS nights (Fig. 3b, e) – regular but chronically short sleepers (low TST SD) accumulate a high number of SS nights while irregular (high TST SD) sleepers are split between short and non-short nights.

In contrast, when using the personalized definition of short sleep (pSS), the association between proportion of SS nights and sleep variability rose in magnitude to 0.88, and for SRI to −0.64 (Fig. 3c, f). This suggests that pSS reflects the combination of duration and regularity.

### Recovery sleep characteristics

A single night of pSS was followed by an increase of approximately 50 minutes in recovery sleep duration on the subsequent night, if it fell on a weekday (Fig. 6d, blue bars). However, additional consecutive nights below the pSS threshold were associated with only modest further increases in recovery sleep on weekdays, amounting to less than 10 minutes per additional night of short sleep. This pattern suggests that weekday recovery sleep is constrained. Note that the data here relates only to the first night following last short sleep night in the run.

**Fig. 6.**
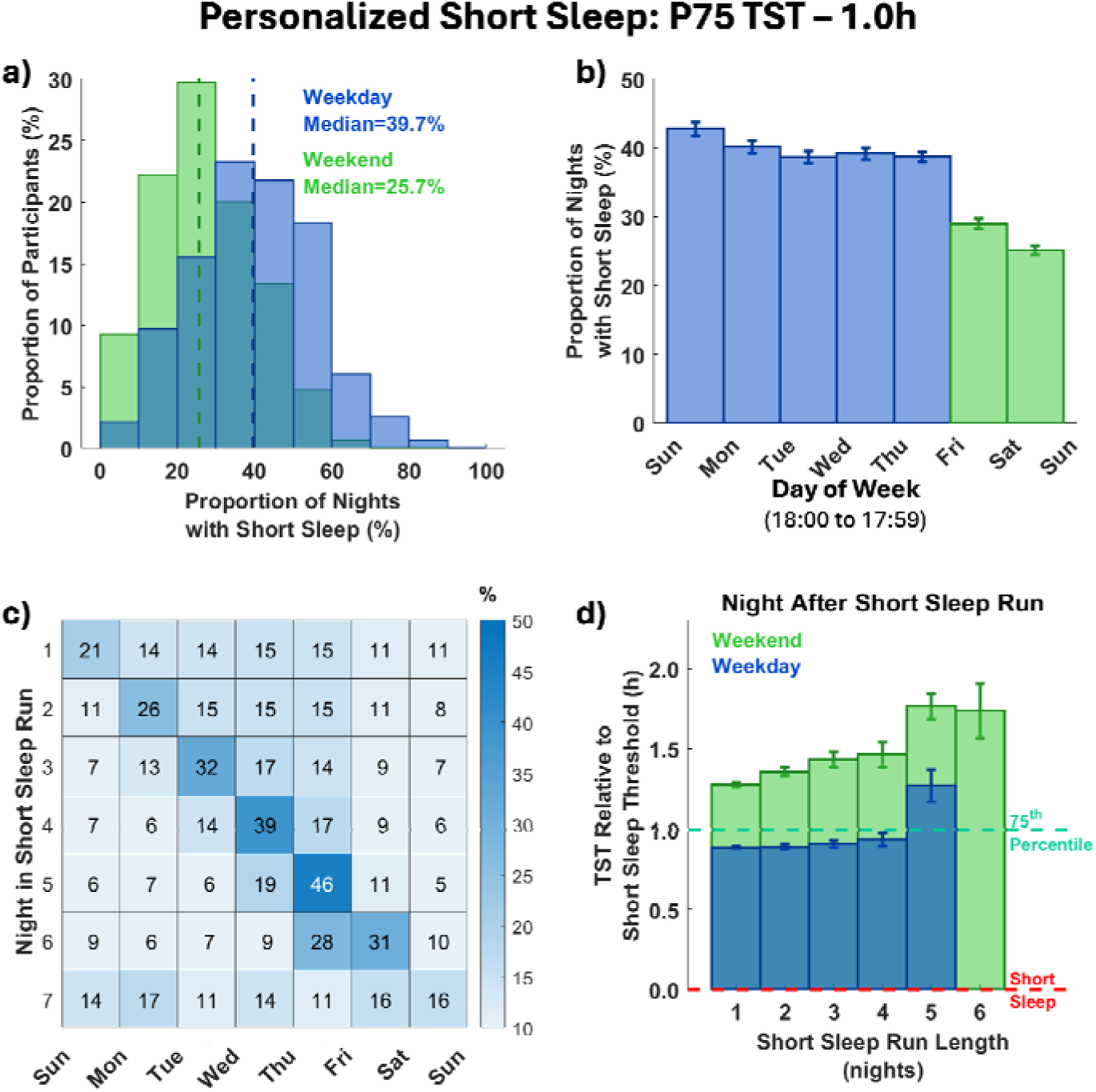
Profile of pSS across the week. (a) and (b): The proportion of pSS was higher on weekdays than on weekends. (c) The night within a pSS run corresponding to the day of the week, e.g., the 5^th^ night within a SS run had a 46% chance of falling on Thursday night. Weekends were a barrier to a run of SS nights. (d) On a weekday night after a short sleep run, participants obtained recovery sleep below their own P75 TST (1.0h longer than pSS threshold). On weekends recovery sleep exceeded P75 by an average of 0.76 [0.08] h (mean [S.E.M.]). Weekdays are shown in blue and weekends in green.

When recovery sleep occurred on weekends, greater sleep opportunity was associated with a larger rebound, with recovery sleep increasing by up to approximately 1.25 hours. However, even on weekends, the increment in recovery sleep following consecutive nights of pSS did not exceed 10 minutes after four successive short-sleep nights. This plateau suggests that recovery sleep following accumulated inadequate sleep is incomplete reflecting interacting social and physiological constraints.

### Short sleep in relation to day-of-week

Nights of short sleep are not randomly distributed. The proportion of weekdays with pSS (median [IQR] = 39.7% [21.4%]) was higher (*t_463_*= 11.0; *p* < 10^−3^) than the proportion of weekends with pSS (median [IQR] = 25.7% [18.6%]; Fig. 6a). Sunday night was most likely to be short; Saturday night was most likely to be long (Fig. 6b). Unsurprisingly, the weekend serves as a barrier to a run of SS nights.

### Reproducibility of sleep findings across multiple countries

While the overall prevalence of pSS and typical run lengths varied systematically across populations, the dominant pattern of isolated short-sleep nights and progressively rarer longer runs of ‘short sleep’ was preserved. Countries with earlier and longer sleep, such as Australia, exhibited lower proportion of pSS nights and shorter run lengths (χ²=4156.7, df=36, p<0.001: Fig S5), whereas countries with later and shorter sleep, such as Japan, showed higher proportion of pSS nights and a higher mean maximum run length (Australia: 3.94; Japan: 4.43; z=17.8, p<0.001: Fig S6 and Table S12).

Across countries, weekday recovery following short-sleep runs rises modestly but generally remains below or near P75 and is often <1 h above the individualized threshold (Fig S6). Weekend recovery, by contrast, is clearly larger and increases with run length, but does not scale linearly. This nonlinearity is noteworthy. If recovery reflected unconstrained or approximately proportional repayment of accumulated sleep loss, longer runs of short sleep might be expected to produce proportionally larger recovery episodes. Instead, we observed modest early recovery, diminishing incremental gain and plateau-like behavior that was strikingly consistent across nations.

## Discussion

In everyday life, isolated nights of short sleep are common, whereas longer runs are relatively infrequent but associated with poorer sleep satisfaction, motivation, sedentariness, and autonomic markers. A personalized definition of short sleep derived from long-term wearable data showed stronger associations with multiple functional markers compared with fixed thresholds. By anchoring sleep deficiency to an individual’s habitual sleep distribution, the proportion of pSS nights, relative sleep shortfall beyond absolute duration, implicitly incorporating information about sleep variability and providing insight into the functional impact of successive short-sleep nights. These distributional patterns generalized across countries differing widely in average sleep duration and sociocultural environment, suggesting that the dynamics of recurrent sleep shortfall may be conserved despite differing sleep practices.

### Motivation and benefits of personalizing the specification of ‘short sleep’

Approximately 7-8 hours of self-reported sleep has consistently been associated with optimal mortality^7^, cognitive^34^, and cardiovascular outcomes^35,36^. However, about a third of individuals worldwide do not meet the current NSF fixed, self-report–based sleep duration recommendations^37^. If wearable device assessed total sleep time is taken at face value, even more will be assessed as having inadequate sleep as objective sleep measurement typically results in lower TST than self-report^10,11^ motivating consideration of a new approach.

Our approach to personalization of short sleep / sleep adequacy is based on three observations. First, individuals are often less concerned with achieving an idealized sleep duration than with obtaining as little sleep as possible without compromising daily functioning. Second, naturalistic sleep recordings provide ecologically valid insight into everyday sleep behavior and its functional consequences^23^. Third, a person’s habitual sleep duration reflects the interaction of sleep need, opportunity, and ability^38,39^, varying around this to respond to life’s demands^23^.

The functional impact of personalizing short sleep is not apparent from average sleep duration alone. At the cohort level, both the mean and median pSS threshold (≈6.1 h) and the overall proportion of nights below threshold were similar to those obtained using fSS, with approximately 36% of nights classified as short under either approach. However, this similarity masked substantial night-level reclassification. Among individuals with shorter pSS thresholds (<6.0 h), fSS classified more nights as short than pSS, whereas longer pSS thresholds identified nights missed by fSS. Thus, similar group-level prevalence concealed meaningful individual-level differences in short-sleep classification. Across countries, populations sleeping less on average also showed higher proportions of pSS crossings. These observations suggest that shorter-sleeping societies experience more recurrent sleep curtailment relative to their own realized sleep distributions rather than simply shorter habitual sleep alone.

pSS discriminates between qualitatively different types of short sleepers and identifies persons appearing to have adequate sleep on average but whose high sleep variability signals sleep shortfall. Two individuals may both average approximately five hours of sleep per night yet differ substantially in their exposure to sleep deficiency. A natural short sleeper or a healthy older adult with diminished sleep capacity, may cross a low pSS threshold infrequently while maintaining short but regular sleep, whereas individuals whose curtailed sleep reflects social pressure, chronic stress, or diminished sleep capacity are more likely to cross this threshold repeatedly and experience recurrent functional impairment (Table 2). Conversely, individuals with few or no nights of <6 h sleep may still show high proportions of pSS if their sleep is highly variable, suggesting that average duration alone may not fully capture sleep adequacy.

**Table 3.**
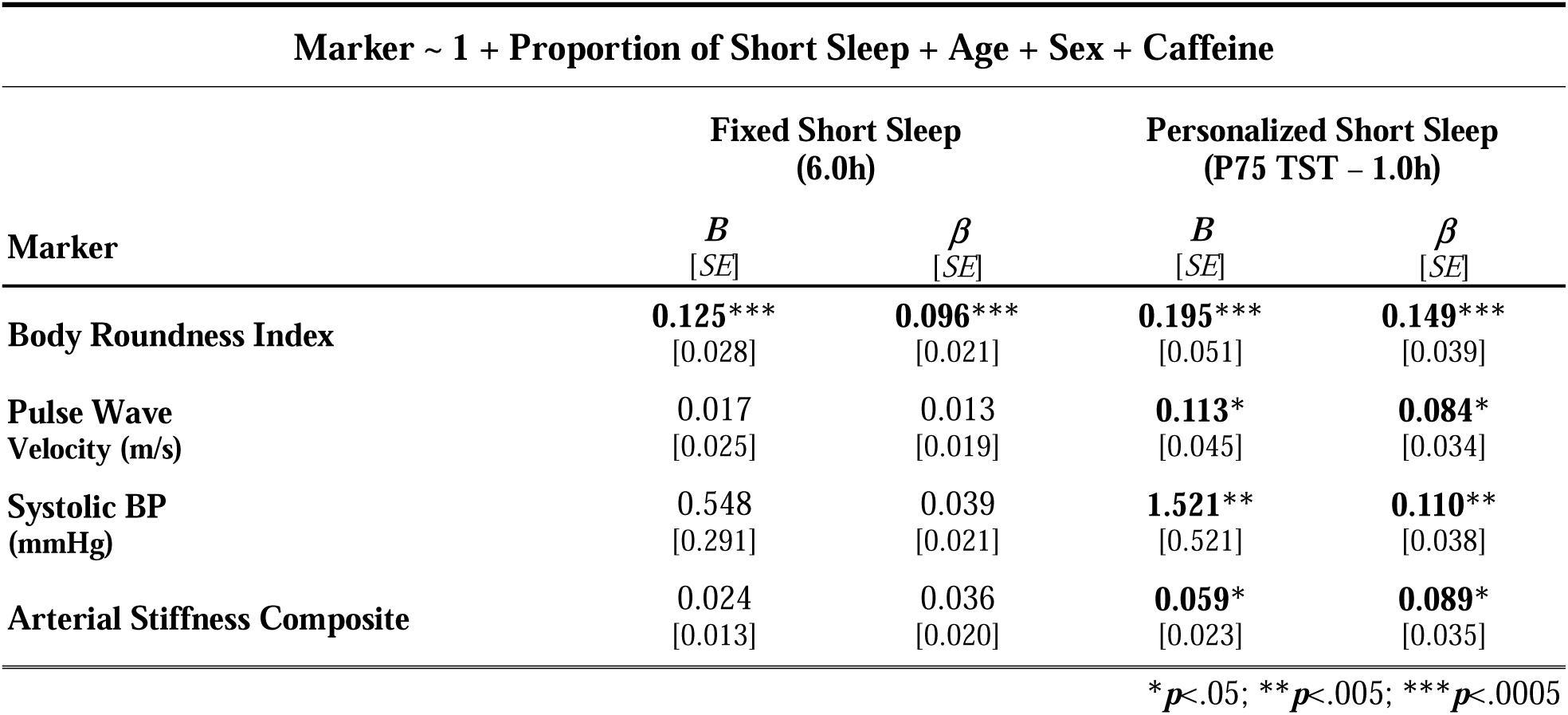
Personalized vs Fixed Definitions of Short Sleep: Associations Between Proportion of Short Sleep and Cardiometabolic Markers.

### Personalizing short sleep improves association with health and wellbeing measures

When modeling night-to-night, accumulative effects of sleep deficiency on short-term, state-sensitive outcomes, pSS consistently showed stronger associations than fixed-duration short sleep (fSS). These outcomes are inherently dynamic and respond to deviations from an individual’s habitual sleep^38^, making them particularly sensitive to the temporal structure of sleep deficiency and recovery. Critically, linking the frequency of pSS crossings to relevant functional and physiological markers alleviates the concern that all short sleepers may be unhealthy^40^.

Similarly longer-term outcomes reflecting cumulative exposure, measures of cardiometabolic risk, personalized definition of short sleep continued to show stronger association.

### Proportion of short sleep nights and sleep variability

Sleep regularity has emerged as a strong predictor of health outcomes, often exceeding sleep duration in predictive power^41–43^. However, sleep duration is more actionable in daily life, as it reflects a recurring decision about time allocation that implicitly involves timing. Sustained improvements in regularity may follow when duration targets are consistently applied.

Our findings indicate that night-to-night variability in sleep duration is dominated by short-sleep deviations rather than symmetric dispersion or extended sleep. Variability is disproportionately influenced by extreme deviations, and in healthy samples such as ours, short nights tend to fall farther from the individual mean than long nights (Fig. S2). This asymmetry may reflect both limits on sleep extension and constraints imposed by social schedules, particularly on weekdays. Consequently, extended sleep contributes relatively little to variability.

### Nights of short sleep occur in isolation or in short runs

Notably, 86.4% of individuals had a median pSS run length of one night, and the maximum run length experienced by an individual averaged four nights. These observations indicate that experimental paradigms involving five or more consecutive nights of sleep restriction^13–15,44,45^—while mechanistically informative—depict relatively uncommon scenarios in daily life^23^. A more ecologically valid approach is therefore to examine the physiological and behavioral consequences of repeated, intermittent deviations from habitual sleep^17^, as captured by personalized pSS thresholds.

### Strengths and Limitations

The key innovation in this work is that instead of explicitly attempting to model sleep need, opportunity, and ability^39^ as separate latent constructs requiring a variety of assumptions including those surrounding sleep homeostasis^23^, we allowed their joint influence to be expressed through each individual’s observed sleep distribution. This distribution-based approach provides a pragmatic form of personalization that is data-adaptive, scalable, and interpretable, while avoiding the complications of more complex modelling strategies.

Our analyses were conducted in generally healthy individuals using a wearable device that has been tested against reference standards for sleep^46^ and heart rate measurement^29^ within our local population.

We found no evidence that attrition or data completeness was driven by short-sleep propensity: the proportion of short-sleep nights was not significantly correlated with the number of days of available data for either fSS or pSS. This argues against survivor bias whereby participants with more frequent short sleep were selectively less likely to contribute longer observation periods.

Although performance characteristics may differ across devices, such differences are unlikely to alter our central conclusions. Applicability to populations with poorer health, in whom sleep assessment may be less reliable, needs to be evaluated.

East Asian populations are known to sleep less than Western populations^21,22^, yet epidemiological data suggest similar relationships between sleep duration and health outcomes^35,36,38^. This observation suggests that cross-cultural differences in realized sleep duration may reflect differences in sleep opportunity and social context rather than intrinsic differences in sleep biology alone^22,38^.

Cardiometabolic markers were assessed at the first study visit, while wearable sleep monitoring began shortly thereafter. Accordingly, the present analyses are associative and do not establish causal directionality. Reverse association remains possible, whereby baseline cardiometabolic status influences subsequent sleep. However, the short interval between assessments and the relative stability of sleep across the year-long monitoring period suggest that the sleep measures captured stable behavioral patterns relevant to the cardiometabolic profile assessed at study entry.

Evening EMA responses may reflect both retrospective evaluation of the previous night’s sleep and influences from same-day mood, fatigue, and workload. This represents an inherent trade-off of single daily EMA protocols designed to minimize participant burden and sustain long-term adherence. The present protocol prioritized ecological validity and year-long compliance while acknowledging that sleep satisfaction and related responses may partly incorporate broader perceptions of daytime functioning.

Finally, seasonal variation and longer-term changes in social obligations could, in principle, influence the computation and application of pSS thresholds. The stability of personalized thresholds across such contextual changes warrants further investigation. However, the effect of seasonal variation on sleep duration is less, even at high latitudes, than the average weekend/weekday difference in sleep duration^47^.

## Conclusion

Taken together, our findings suggest that sleep health may be better characterized by considering recurrent short-sleep exposure relative to an individual’s habitual sleep pattern rather than relying solely on fixed duration thresholds. Consecutive nights of sleep shortfall approximately one hour below habitual sleep patterns were associated with functional effects, whereas isolated short nights were common and typically followed by recovery sleep. Personalized wearable-derived metrics may therefore complement existing sleep recommendations by helping individuals monitor recurrent sleep shortfall without over-interpreting occasional short sleep.

## Acknowledgement

The authors would like to thank Ruth Leong, Gizem Yilmaz, Nicholas Chee, Kian Foong Wong, Liang Tian, Tara Martin, Zhenghao Pu, Alexander Soo, and Annalissa Munoz for their contributions to data collection.

## Data availability

The data in the main study cohort are available upon reasonable request from the authors. However, the multi-country data is not available for sharing.

