## Supplementary Materials for "Personalized Definition of Short Sleep Using Long-Term Wearable Sleep Distributions"

Personalized Definition of Short Sleep Using Distributional Characteristics of Individual Long-Term Wearable Data

### Supplementary Materials

Corresponding author:

Prof. Michael W. L. Chee

Centre for Sleep and Cognition

NUS Yong Loo Lin School of Medicine,

MD1, 12 Science Drive 2

Singapore 117549

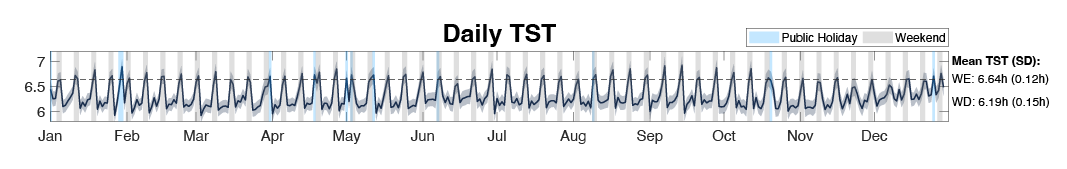

### Supplementary Results

**Fig. S1. Average sleep duration across the year.** To illustrate consistency in the weekly sleep rhythm when participant data is averaged. Weekday sleep duration was ~30 minutes shorter than on weekends / public holidays. Singapore does not have traditional seasons as it lies almost on equator and photoperiod variation is ~10 mins over the year.

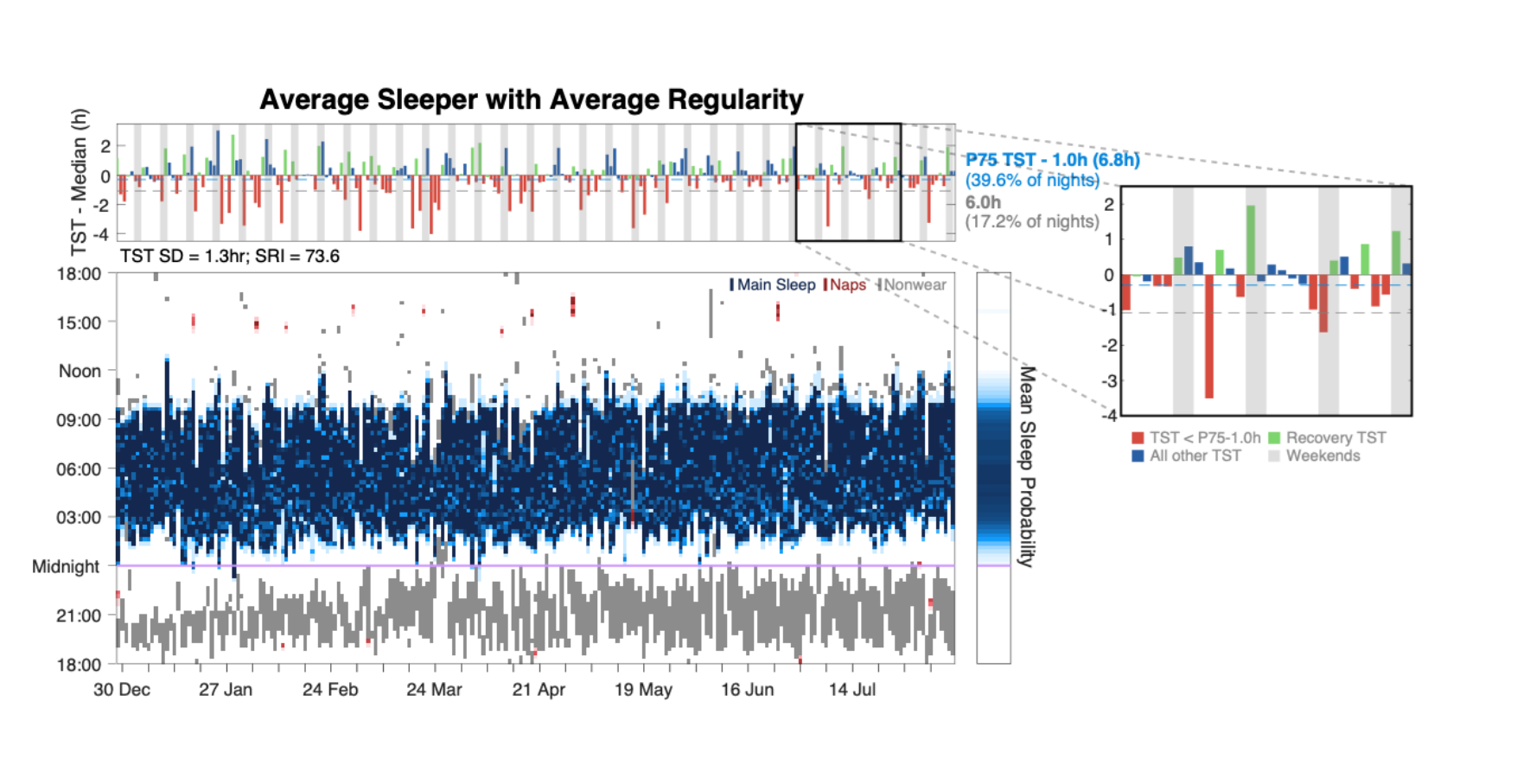

**Fig. S2. Sleep profile of a participant with adequate sleep duration but appears to require more sleep.** Top panel shows daily main sleep duration relative to median total sleep time (TST; 7.1h). In contrast to the participant in Fig. 4, this person experienced fewer nights below the fSS threshold (grey dashed line) but has many more nights below their pSS threshold (blue dashed line, 6.8h) suggesting that this person requires more sleep than the population average to achieve ‘adequacy’. Most pSS nights (red) occurred on weekdays. Bottom panel shows daily main nocturnal sleep (blue), naps (red) and non-wear time (grey). The figure highlights the tendency for SS nights to show larger deviations from habitual TST, compared to recovery nights (green) and nights following the recovery night, that having smaller ‘compensatory’ excursions.

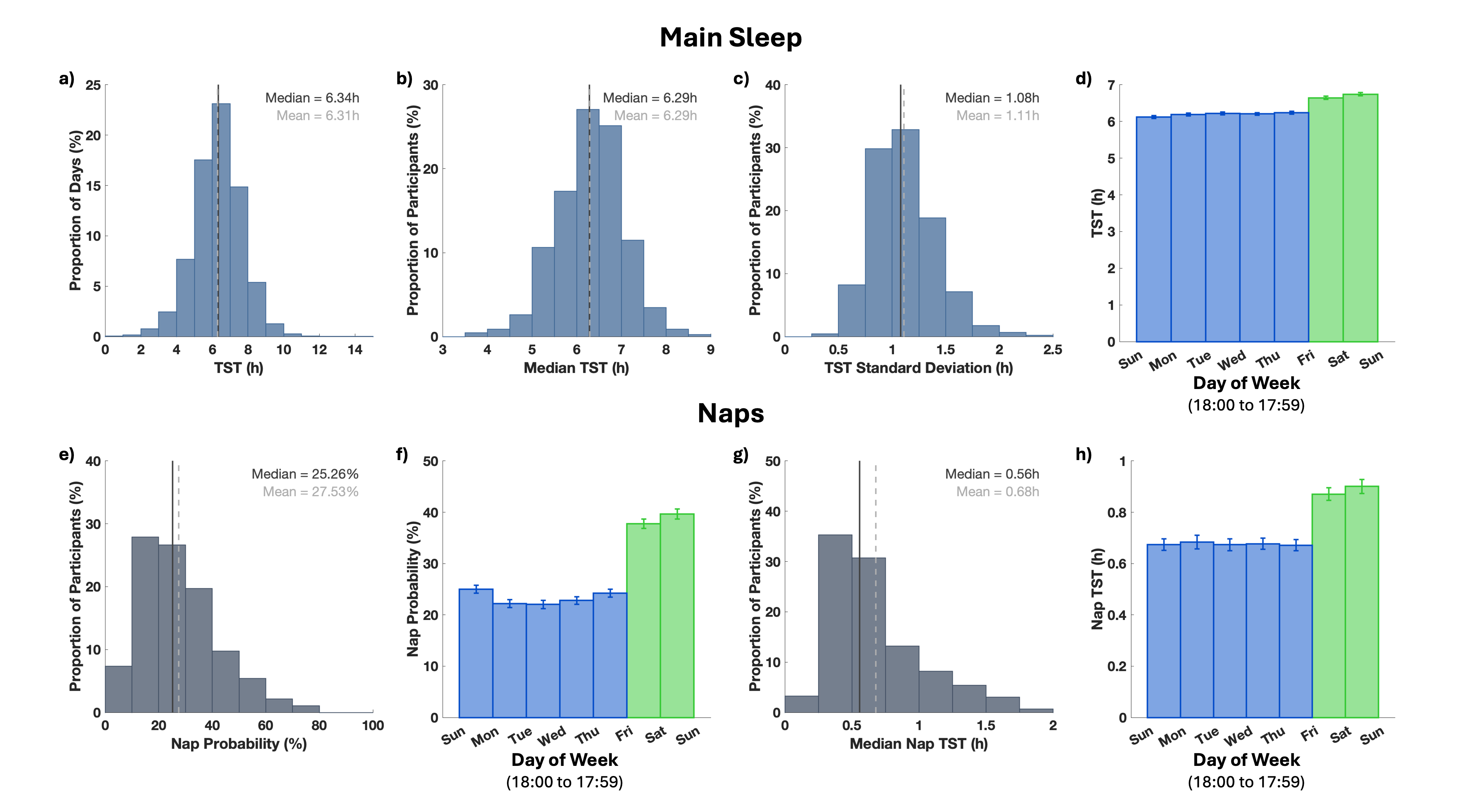

**Fig. S3. Duration and variability of main sleep and naps.** (a) Distribution of main sleep TST across all nights. (b) Distribution of participants’ median main sleep TST. (c) Distribution of within-person standard deviation of main sleep TST. (d) Mean main sleep TST by day of week (mean ± S.E.M. across participants), e.g., first bar represents the mean main sleep duration recorded between Sunday 18:00 to Monday 17:59. (e) Distribution of participant-level nap probability, defined as the percentage of all days (including days without naps) on which at least one nap was recorded. (f) Mean nap probability by day-of-week (mean ± S.E.M. across participants), e.g., the number of days of a given day-of-week when napping occurred divided by the total number of recorded days for that day-of-week. (g) Distribution of participants’ median nap duration, computed only on days when napping occurred. (h) Mean nap TST by day-of-week (mean ± S.E.M. across participants), averaged across all days of a given day-of-week, including days without naps. Weekdays are shown in blue and weekends in green.

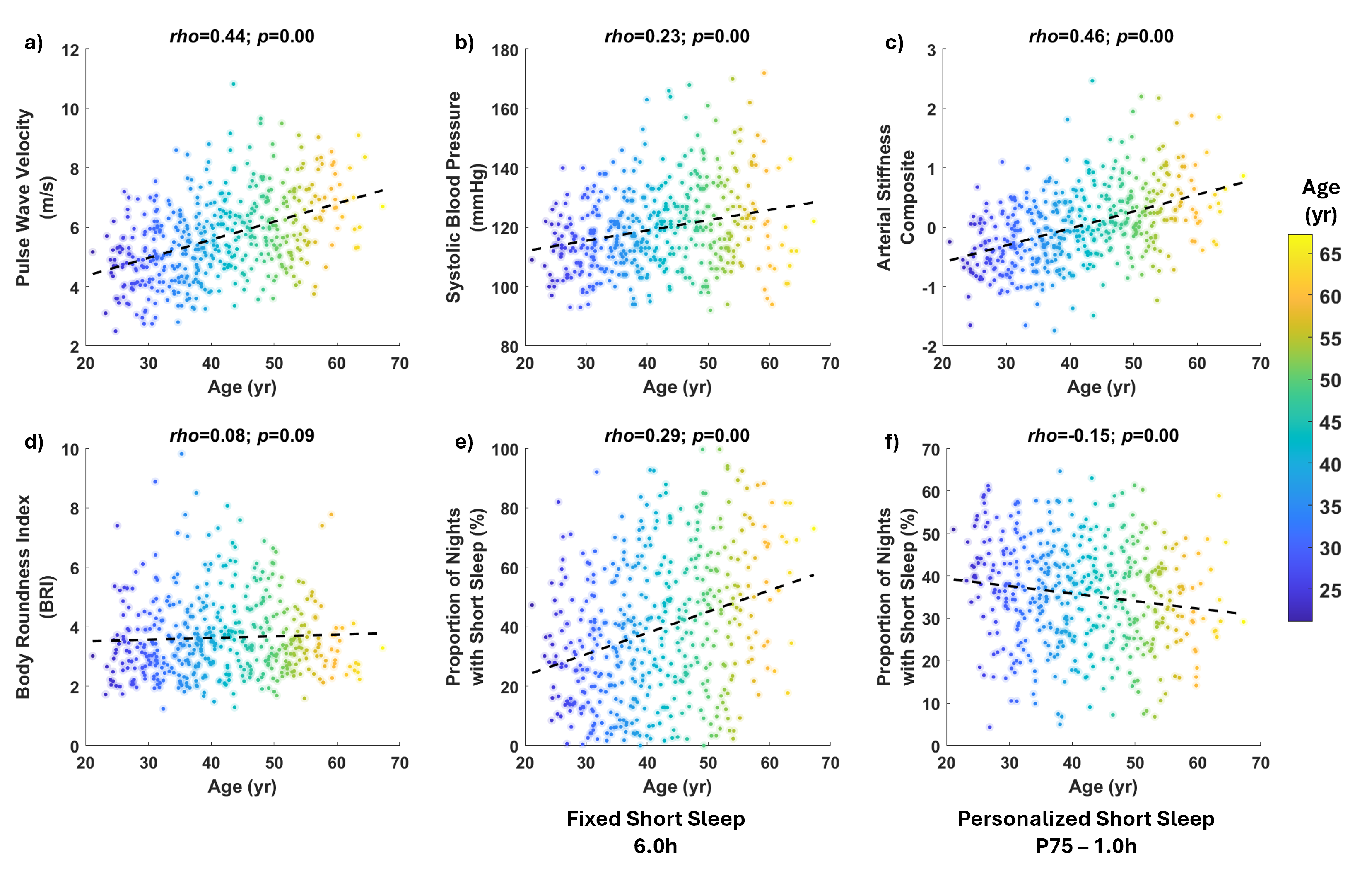

**Fig. S4. Age was correlated with various cardiovascular health indicators and proportion of SS.** Of the cardiovascular health indicators, only BRI was not associated with age. With increasing age, the proportion of fSS also increased while the opposite trend was observed for pSS. This underscores the importance of controlling for age in the LMMs.

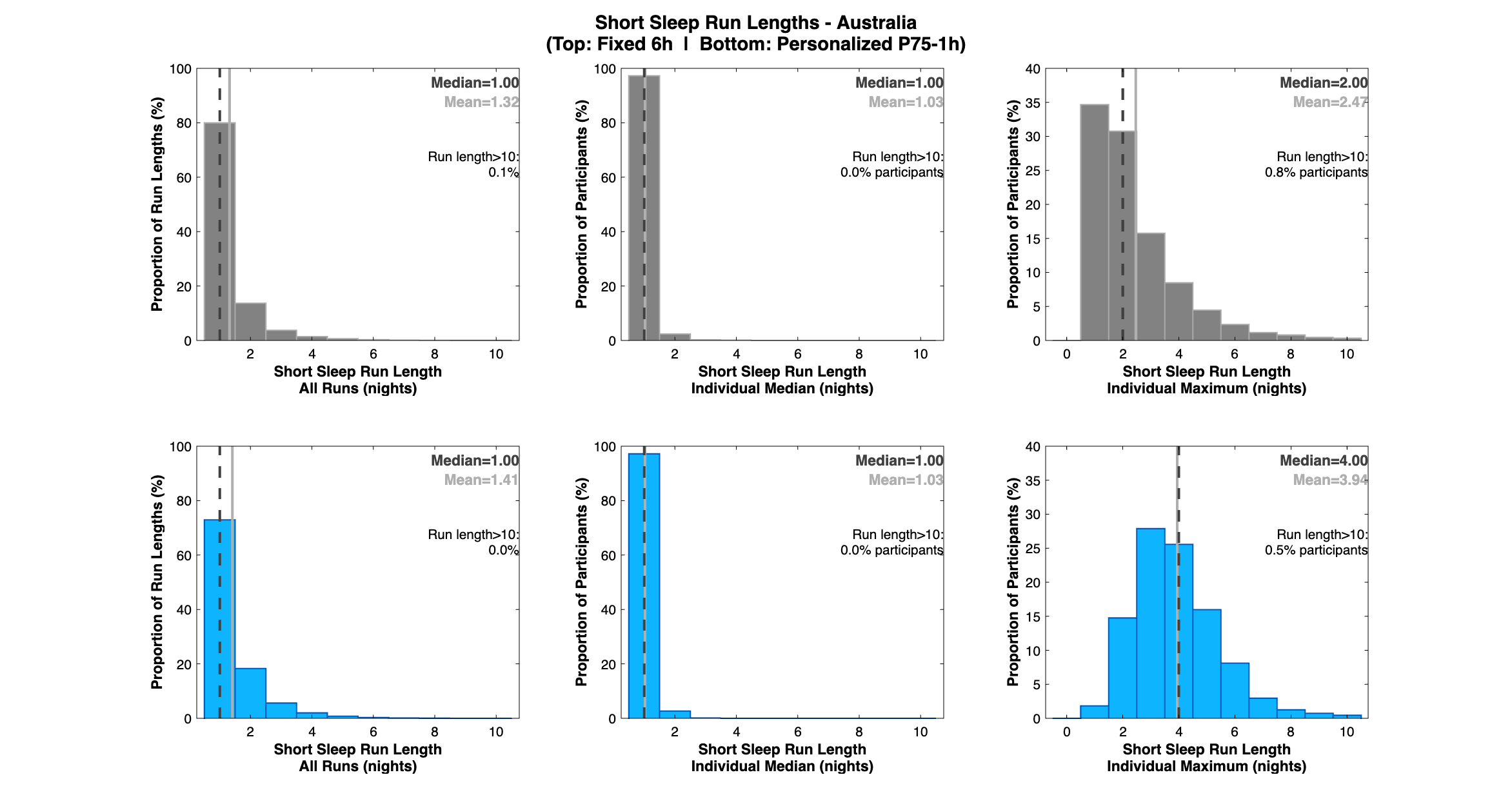

**Fig. S5. fSS and pSS run length distributions for Australia.** Refer to Fig. 2 captions for descriptions of each panel.

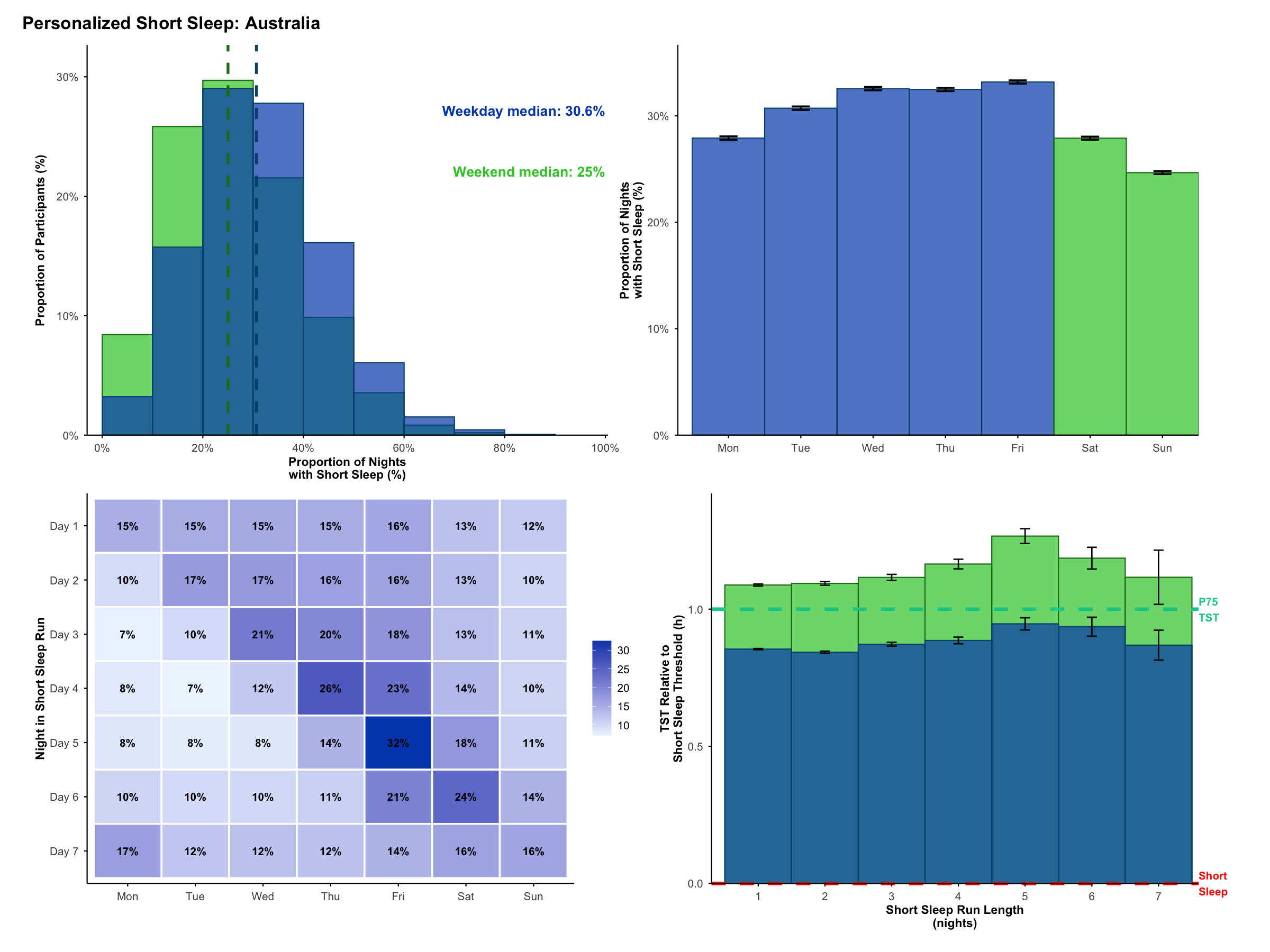

**Fig. S6. Profile of pSS across the week for Australia.** Refer to Fig. 6 captions for descriptions of each panel.

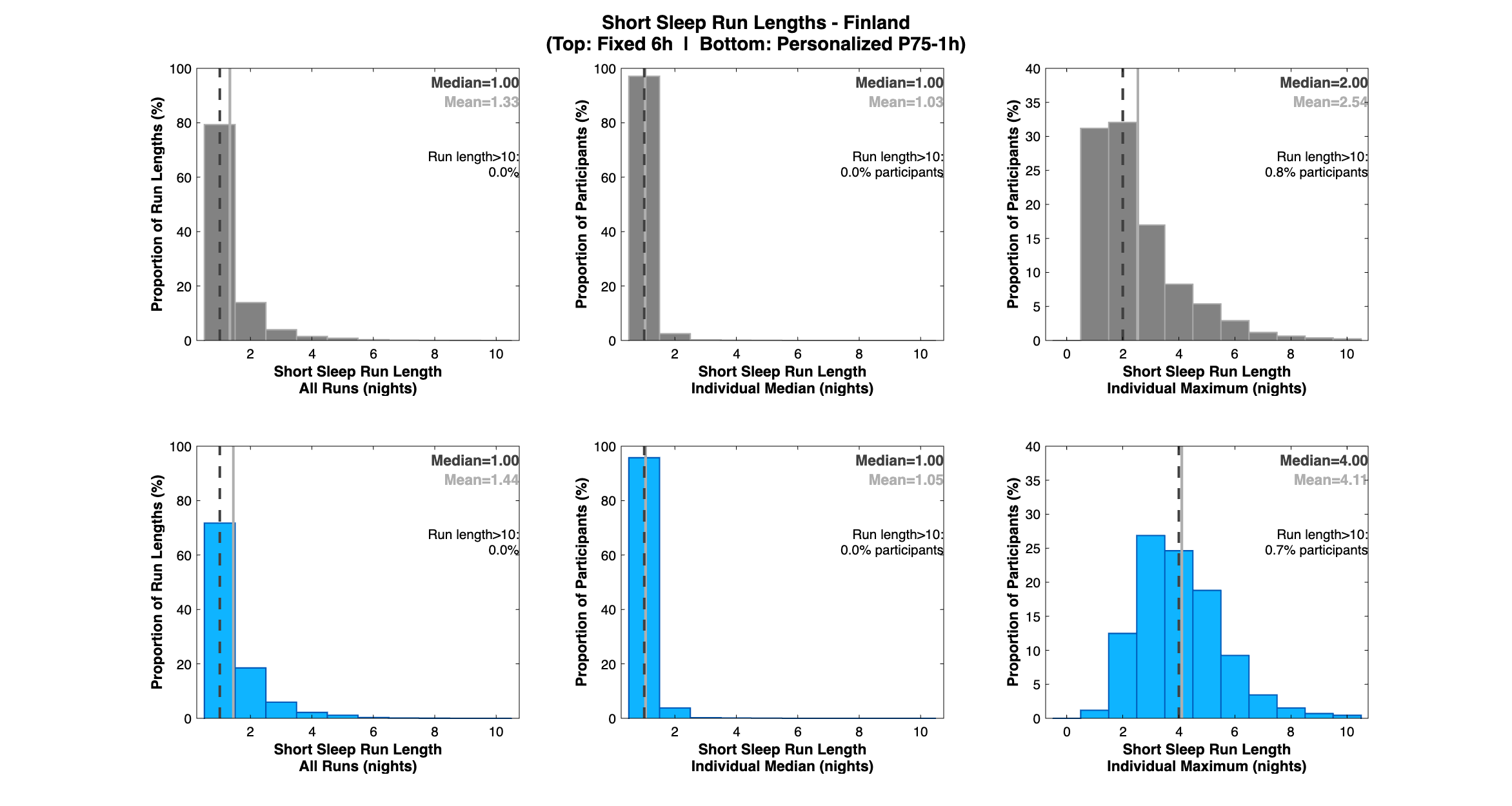

**Fig. S7. fSS and pSS run length distributions for Finland.** Refer to Fig. 2 captions for descriptions of each panel.

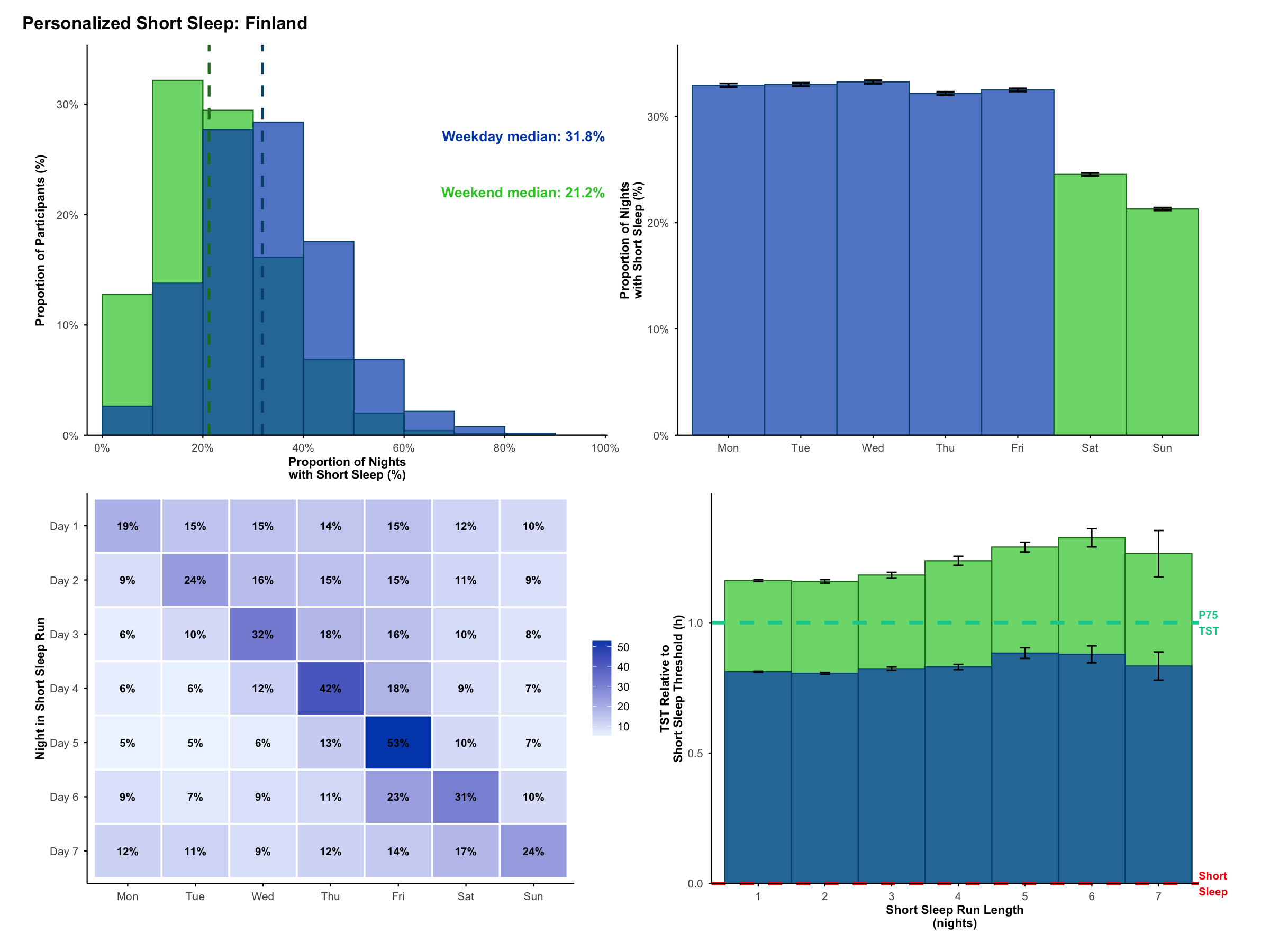

**Fig. S8. Profile of pSS across the week for Finland.** Refer to Fig. 6 captions for descriptions of each panel.

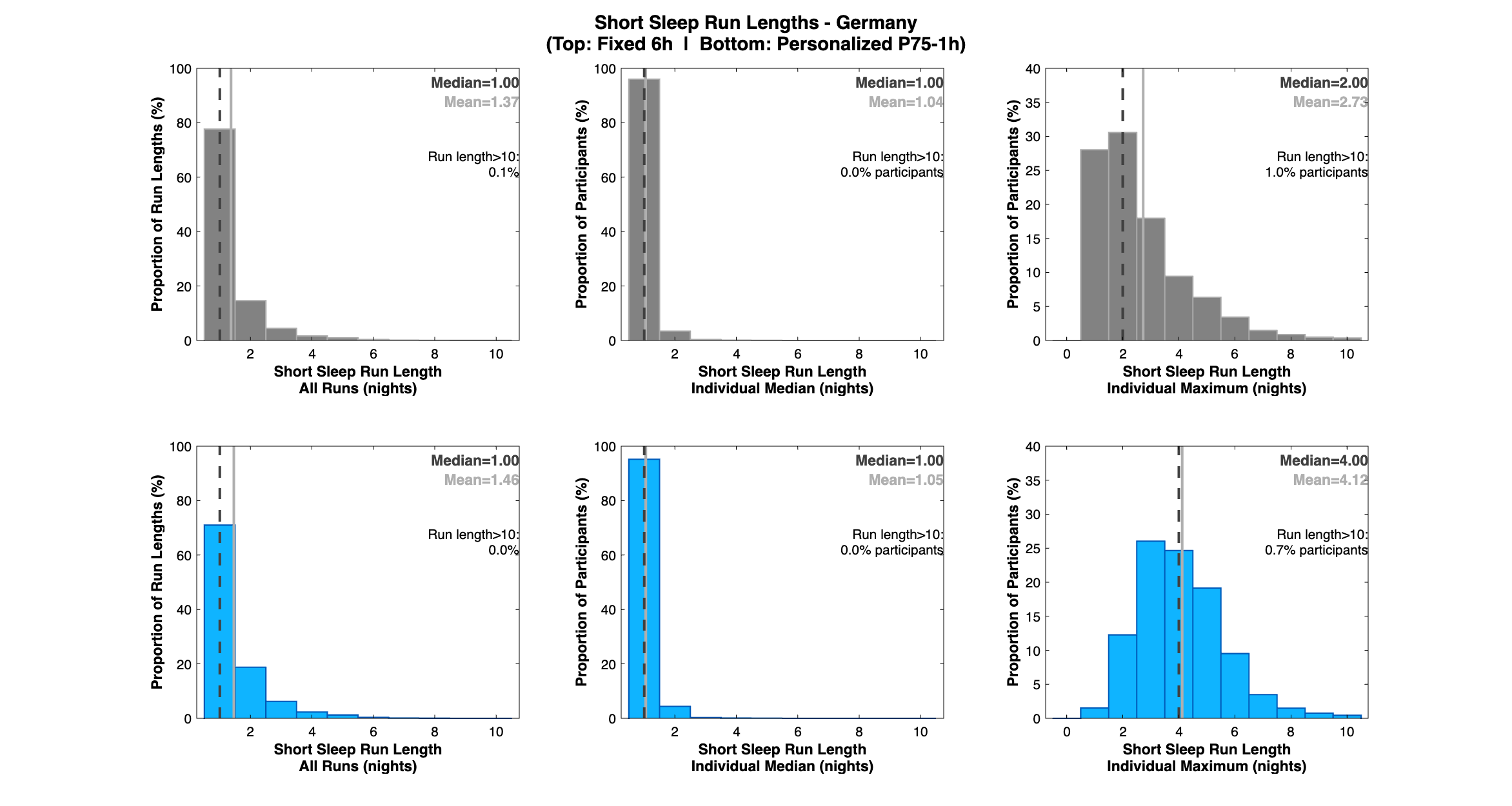

**Fig. S9. fSS and pSS run length distributions for Germany.** Refer to Fig. 2 captions for descriptions of each panel.

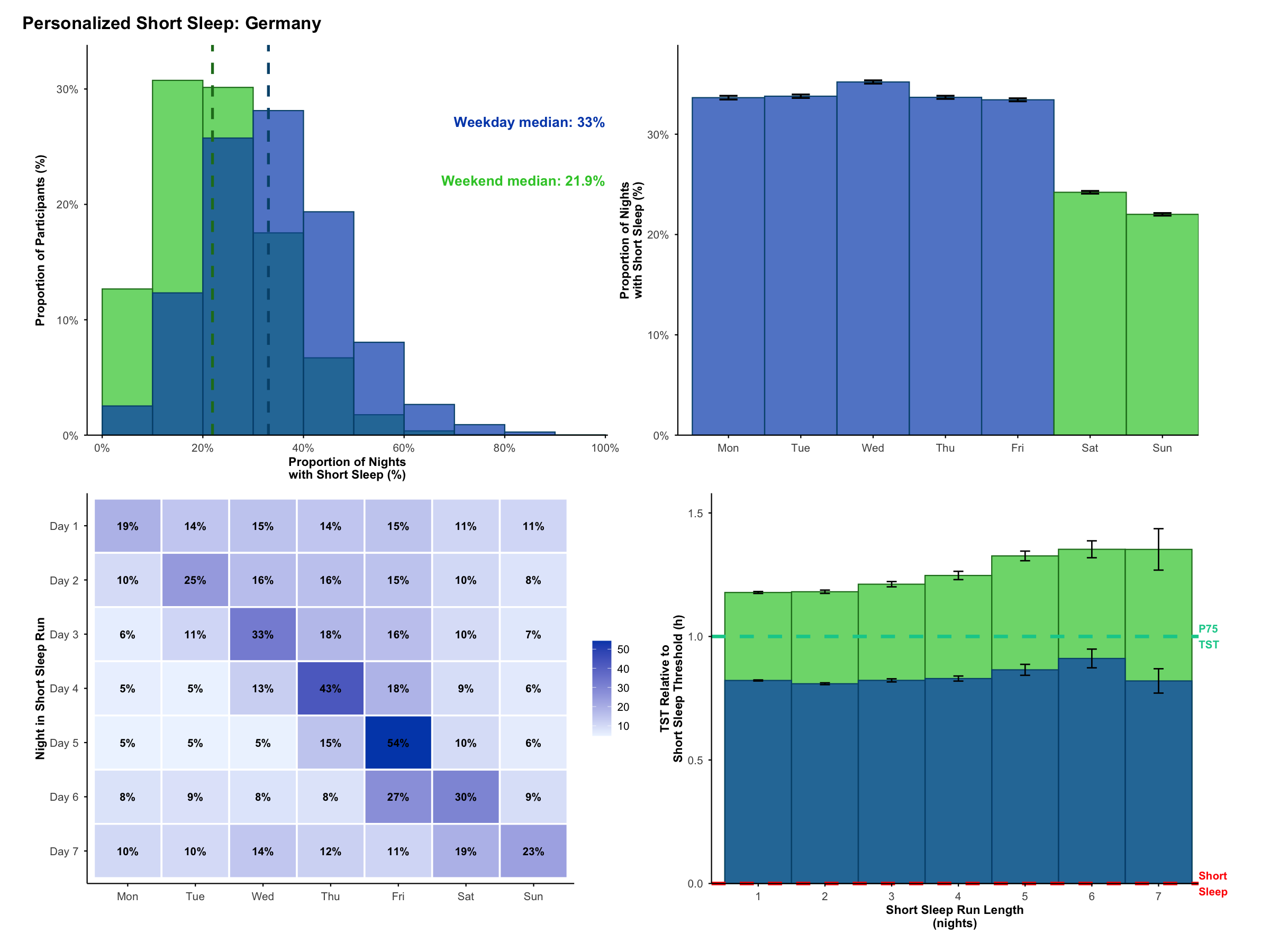

**Fig. S10. Profile of pSS across the week for Germany.** Refer to Fig. 6 captions for descriptions of each panel.

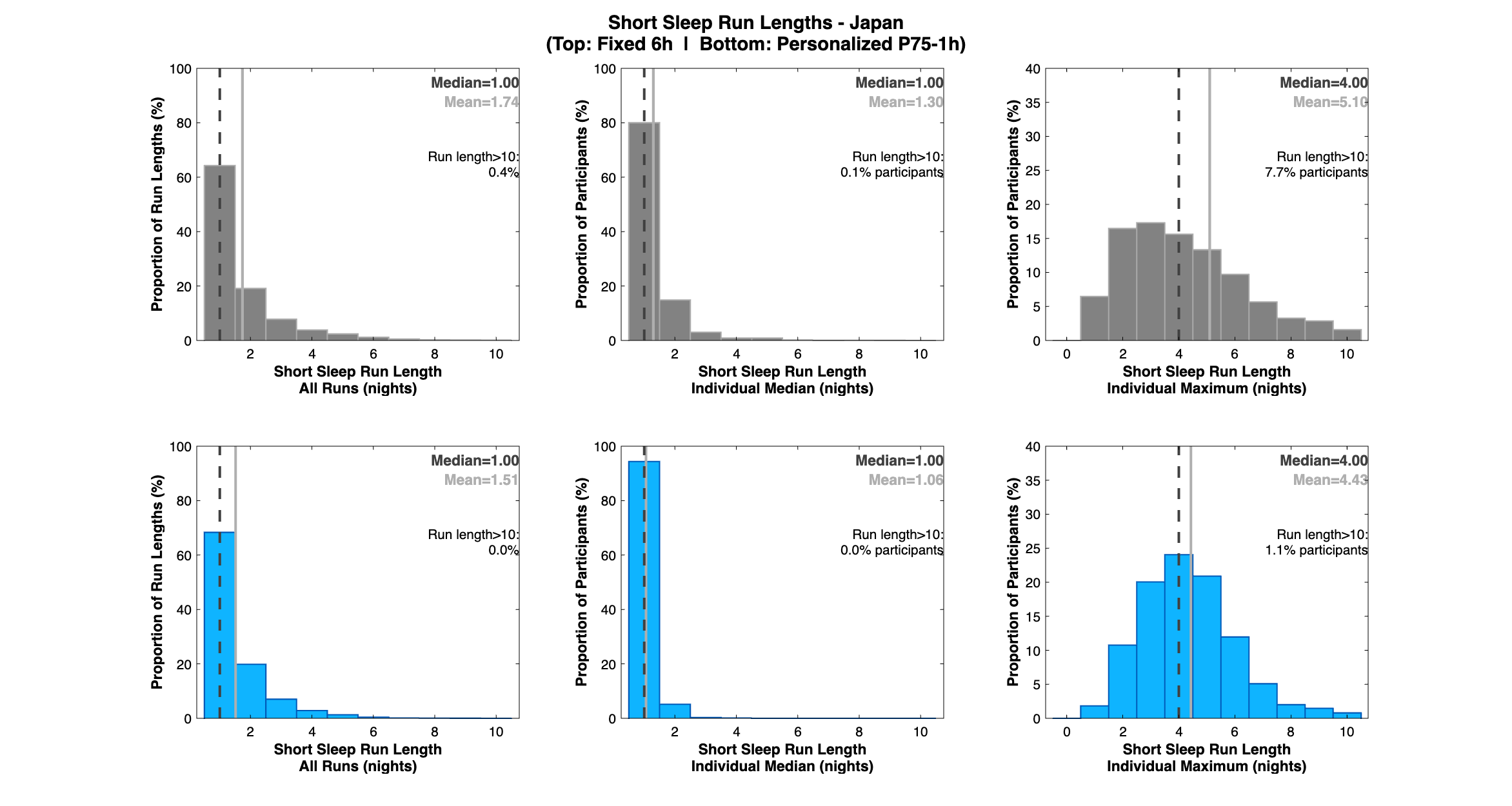

**Fig. S11. fSS and pSS run length distributions for Japan.** Refer to Fig. 2 captions for descriptions of each panel.

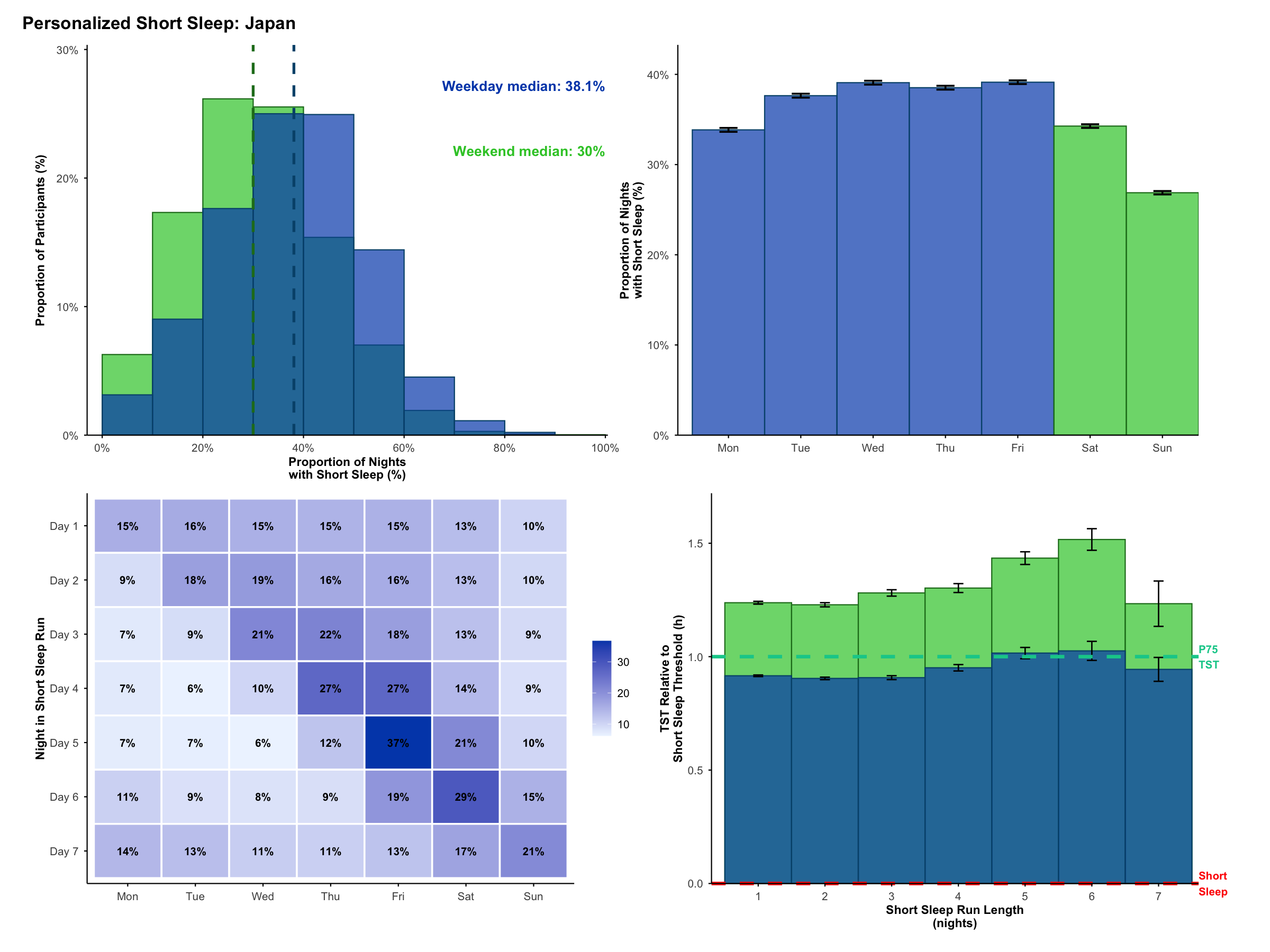

**Fig. S12. Profile of pSS across the week for Japan.** Refer to Fig. 6 captions for descriptions of each panel.

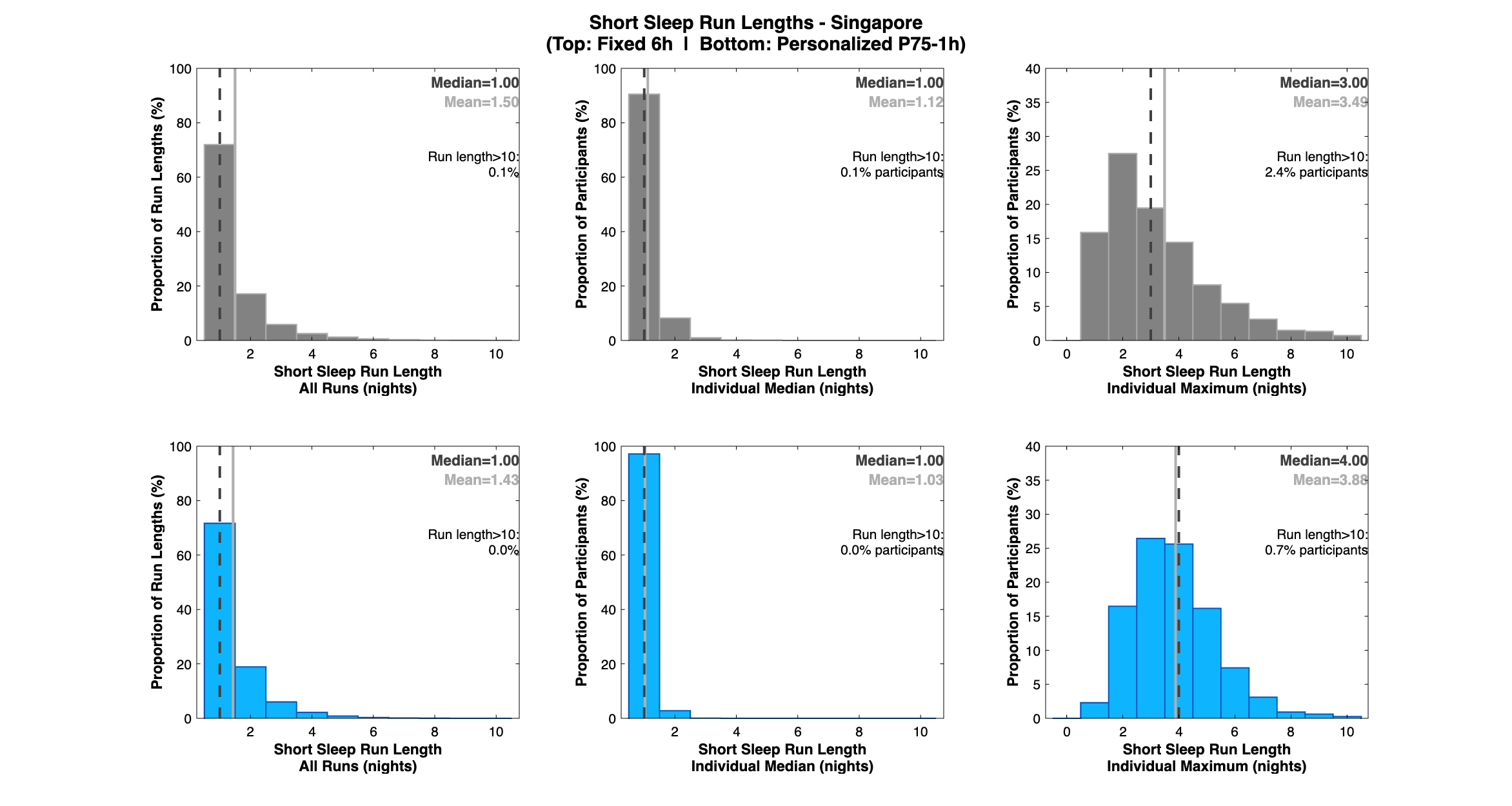

**Fig. S13. fSS and pSS run length distributions for Singapore.** Refer to Fig. 2 captions for descriptions of each panel.

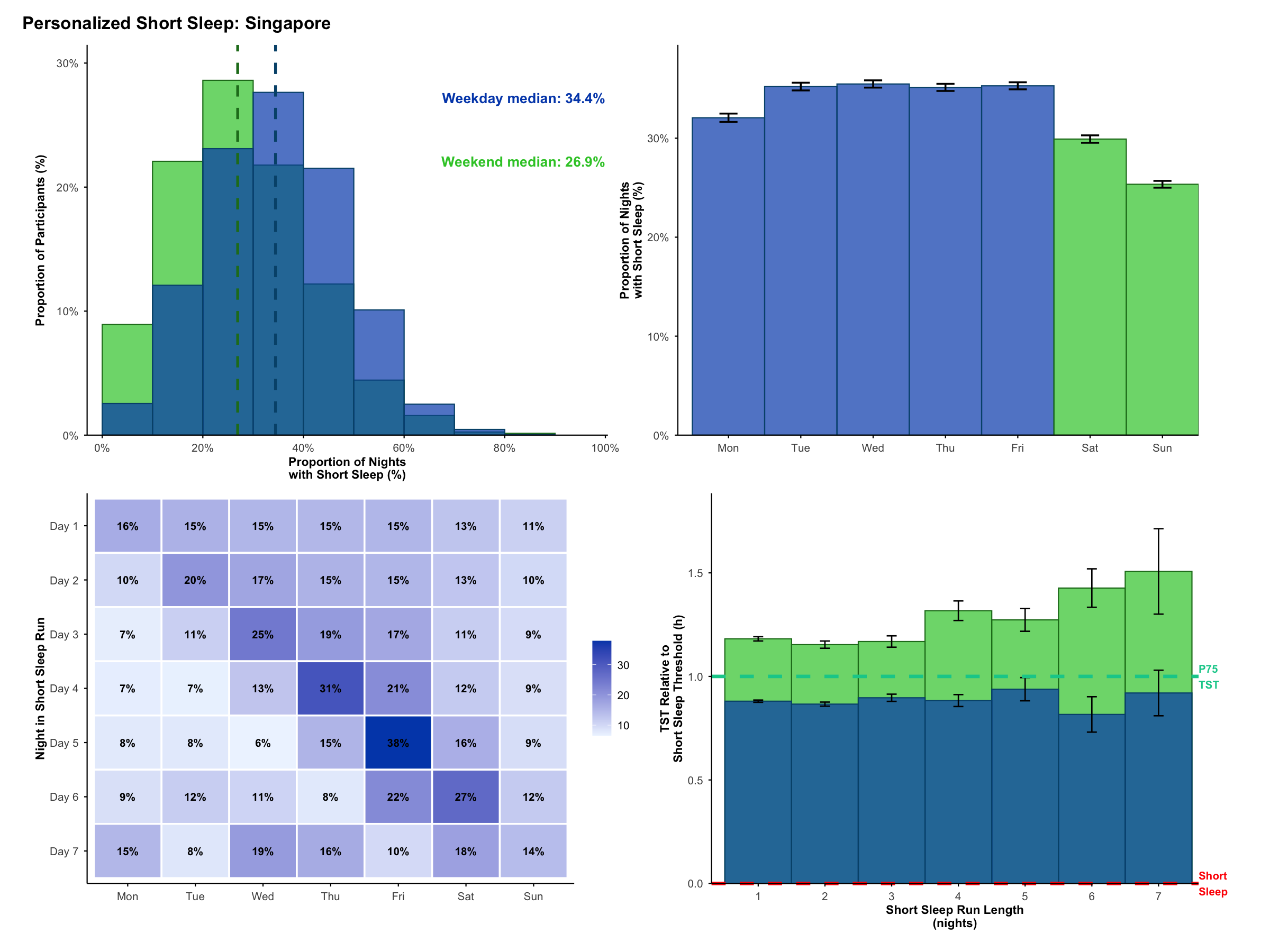

**Fig. S14. Profile of pSS across the week for Singapore.** Refer to Fig. 6 captions for descriptions of each panel.

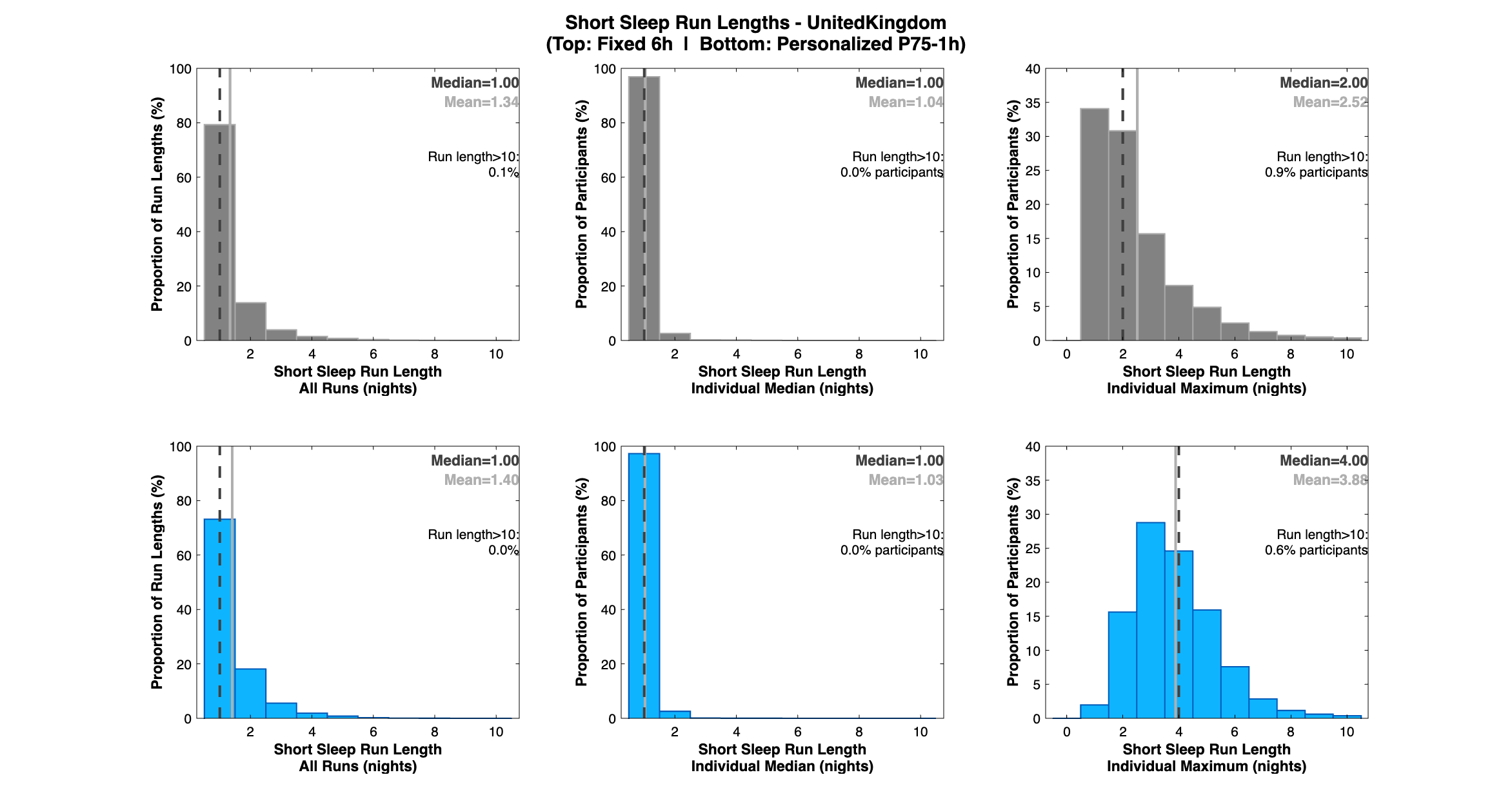

**Fig. S15. fSS and pSS run length distributions for United Kingdom.** Refer to Fig. 2 captions for descriptions of each panel.

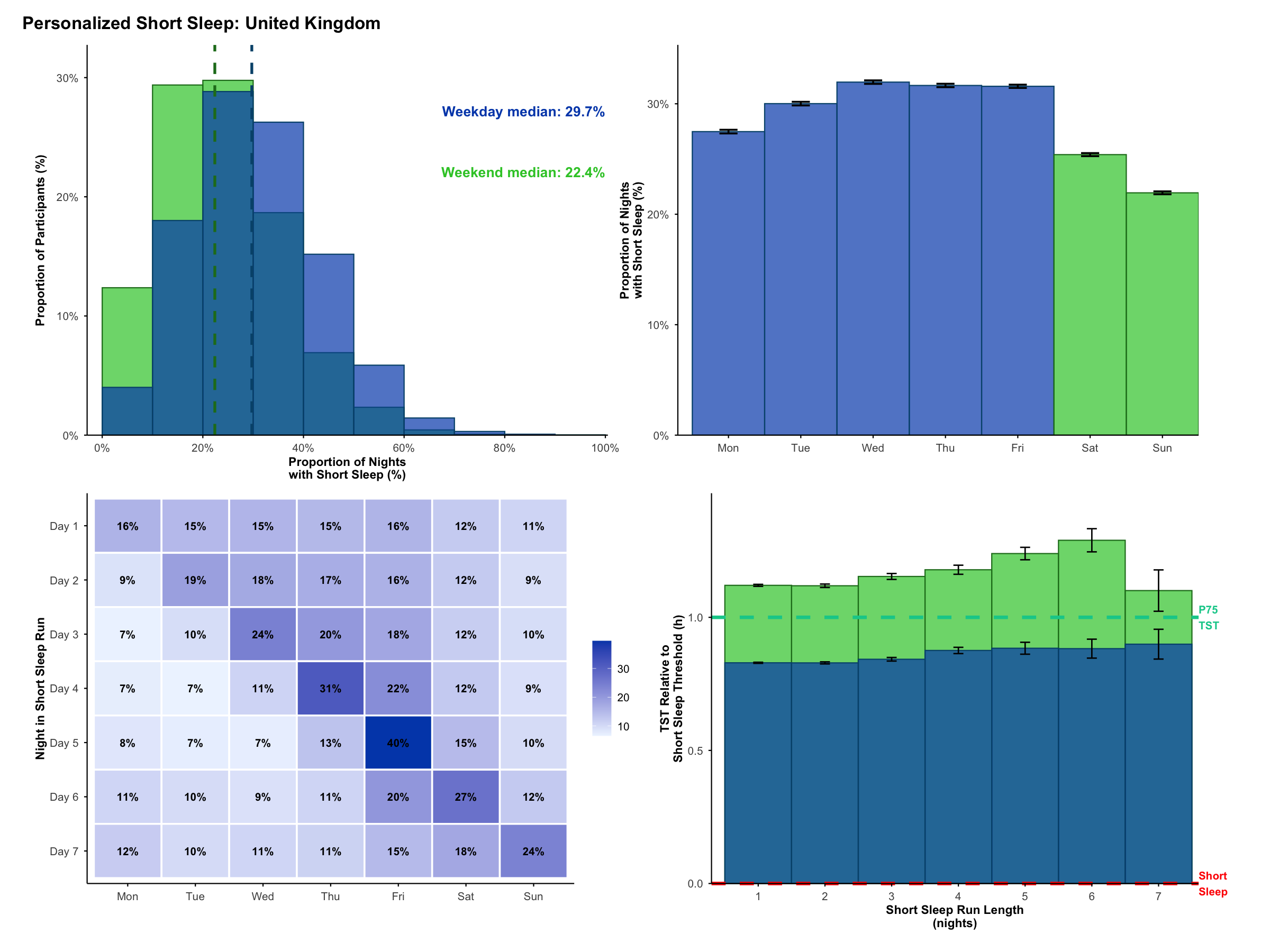

**Fig. S16. Profile of pSS across the week for United Kingdom.** Refer to Fig. 6 captions for descriptions of each panel.

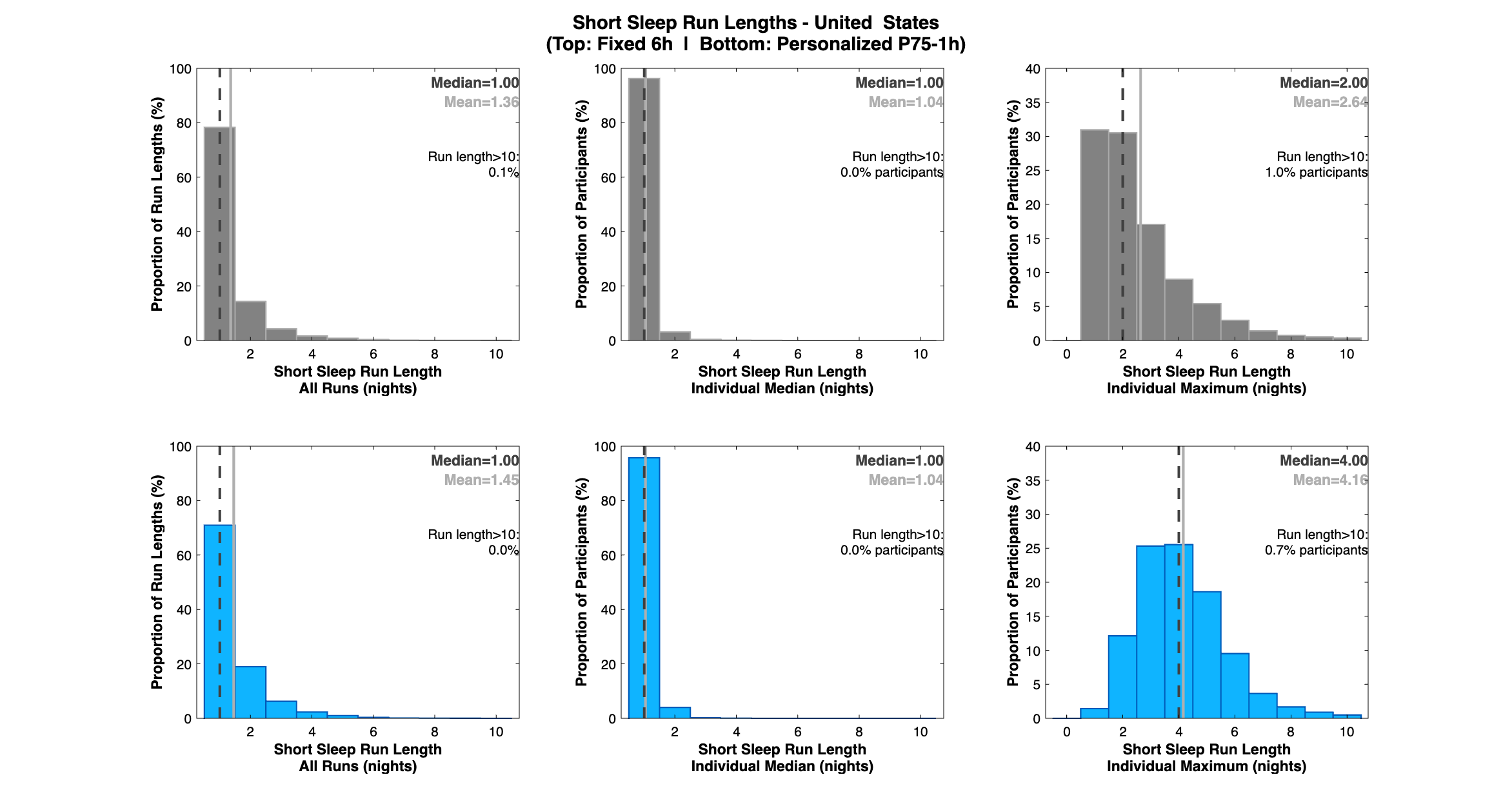

**Fig. S17. fSS and pSS run length distributions for United States.** Refer to Fig. 2 captions for descriptions of each panel.

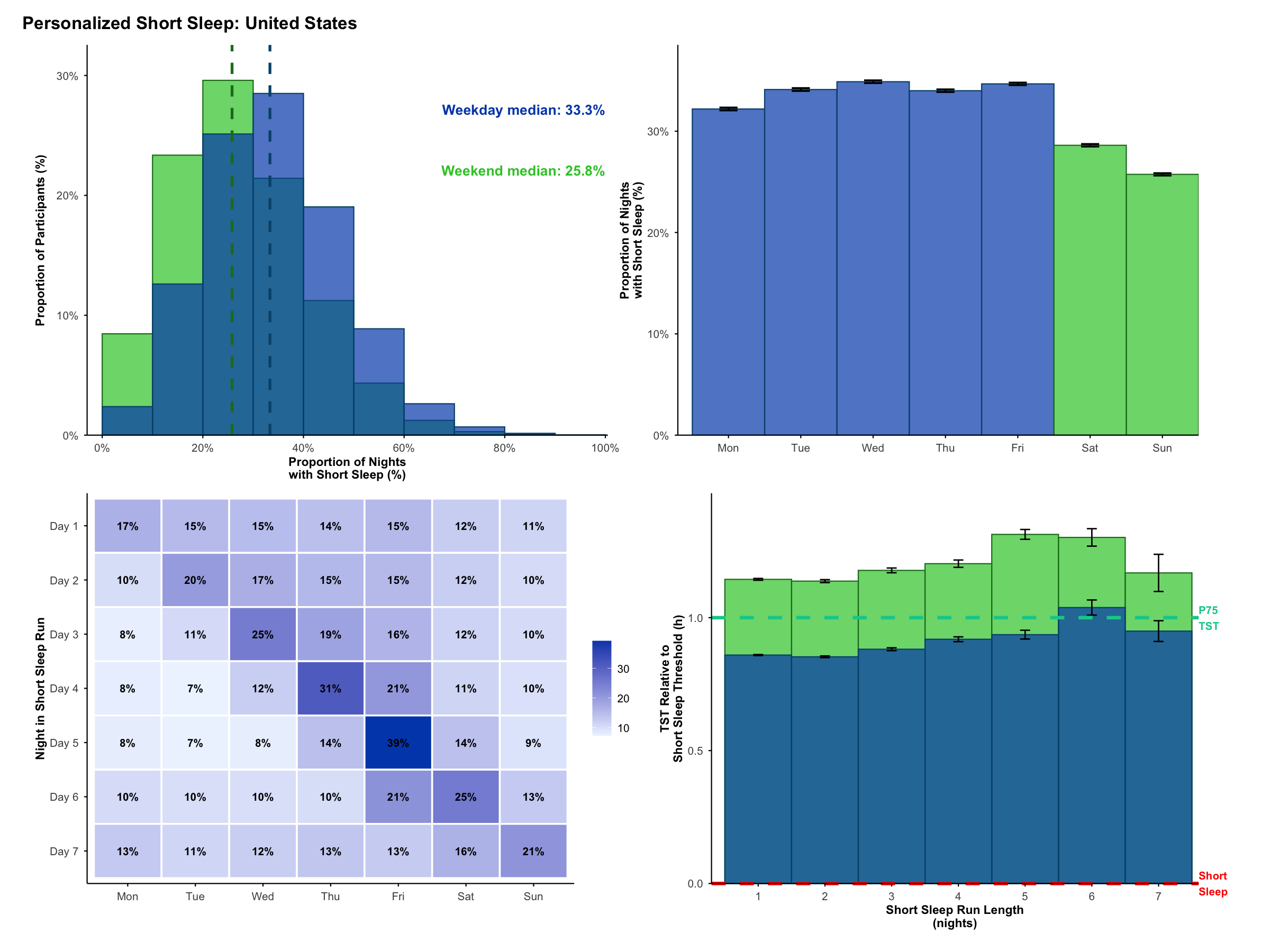

**Fig. S18. Profile of pSS across the week for United States.** Refer to Fig. 6 captions for descriptions of each panel.

#### Table S1. Personalized vs Fixed Definitions of Short Sleep: LMM without Age, Sex and Caffeine Covariates

| **Marker ~ 1 + Night in Sequence + (1\|Subject) + (1\|Subject:Sequence ID)** | | | | | |
| --- | --- | --- | --- | --- | --- |
|  |  | **Fixed Short Sleep (6.0h)** | | **Personalized Short Sleep (P75 TST – 1.0h)** | |
| **Category** | **Marker** | ***B*** [*SE*] | ***β*** [*SE*] | ***B*** [*SE*] | ***β*** [*SE*] |
| ***Mental Wellbeing Markers*** | **Stress** | **0.201***** | **0.008***** | **0.808***** | **0.033***** |
|  | **(Range: 0-100)** | [0.028] | [0.001] | [0.062] | [0.003] |
|  | **Motivation** | **-0.327***** | **-0.012***** | **-0.590***** | **-0.022***** |
|  | **(Range: 0-100)** | [0.025] | [0.001] | [0.055] | [0.002] |
|  | **Sadness** | **0.064**** | **0.004**** | **0.223***** | **0.013***** |
|  | **(Range: 0-100)** | [0.020] | [0.001] | [0.046] | [0.003] |
| ***Sleep Markers*** | **Sleep Satisfaction** | **-0.022***** | **-0.023***** | **-0.119***** | **-0.123***** |
|  | **(Range: 1-5)** | [0.001] | [0.001] | [0.003] | [0.003] |
|  | **Alertness** | **-0.421***** | **-0.017***** | **-1.029***** | **-0.041***** |
|  | **(Range: 0-100)** | [0.022] | [0.001] | [0.048] | [0.002] |
| ***Cardiovascular Markers*** | **Sleep Heartrate** | **0.026***** | **0.003***** | **0.175***** | **0.021***** |
|  | **(bpm)** | [0.006] | [0.001] | [0.013] | [0.002] |
|  | **Heartrate Variability** | **-0.057***** | **-0.003***** | **-0.343***** | **-0.016***** |
|  | **(rMSSD; ms)** | [0.015] | [0.001] | [0.034] | [0.002] |
| ***Physical Activity Markers*** | **Sedentary** | **1.269***** | **0.010***** | **3.645***** | **0.028***** |
|  | **(min)** | [0.170] | [0.001] | [0.378] | [0.003] |
|  | **Low Physical Activity** | **1.194***** | **0.013***** | **4.888***** | **0.051***** |
|  | **(min)** | [0.116] | [0.001] | [0.260] | [0.003] |
|  | **Moderate to Vigorous** | **0.384***** | **0.008***** | **1.345***** | **0.029***** |
|  | **Physical Activity (min)** | [0.056] | [0.001] | [0.124] | [0.003] |
| ****p***<.05; *****p***<.005; ******p***<.0005 | | | | | |

The associations between consecutive days of short sleep and wellbeing indicators were similar to results presented in Table 2, when the Age, Sex and Caffeine covariates were removed. The pSS model was preferred for all indicators.

#### Table S2. Personalized vs Fixed Definitions of Short Sleep: LMM without Caffeine as Covariate

| **Marker ~ 1 + Night in Sequence + Age + Sex + (1\|Subject) + (1\|Subject:Sequence ID)** | | | | | |
| --- | --- | --- | --- | --- | --- |
|  |  | **Fixed Short Sleep (6.0h)** | | **Personalized Short Sleep (P75 TST – 1.0h)** | |
| **Category** | **Marker** | ***B*** [*SE*] | ***β*** [*SE*] | ***B*** [*SE*] | ***β*** [*SE*] |
| ***Mental Wellbeing Markers*** | **Stress** | **0.201***** | **0.008***** | **0.807***** | **0.033***** |
|  | **(Range: 0-100)** | [0.028] | [0.001] | [0.062] | [0.003] |
|  | **Motivation** | **-0.328***** | **-0.012***** | **-0.590***** | **-0.022***** |
|  | **(Range: 0-100)** | [0.025] | [0.001] | [0.055] | [0.002] |
|  | **Sadness** | **0.065**** | **0.004**** | **0.222***** | **0.013***** |
|  | **(Range: 0-100)** | [0.020] | [0.001] | [0.046] | [0.003] |
| ***Sleep Markers*** | **Sleep Satisfaction** | **-0.022***** | **-0.023***** | **-0.118***** | **-0.123***** |
|  | **(Range: 1-5)** | [0.001] | [0.001] | [0.003] | [0.003] |
|  | **Alertness** | **-0.421***** | **-0.017***** | **-1.028***** | **-0.041***** |
|  | **(Range: 0-100)** | [0.022] | [0.001] | [0.048] | [0.002] |
| ***Cardiovascular Markers*** | **Sleep Heartrate** | **0.026***** | **0.003***** | **0.175***** | **0.021***** |
|  | **(bpm)** | [0.006] | [0.001] | [0.013] | [0.002] |
|  | **Heartrate Variability** | **-0.057***** | **-0.003***** | **-0.343***** | **-0.016***** |
|  | **(rMSSD; ms)** | [0.015] | [0.001] | [0.034] | [0.002] |
| ***Physical Activity Markers*** | **Sedentary** | **1.266***** | **0.010***** | **3.651***** | **0.029***** |
|  | **(min)** | [0.170] | [0.001] | [0.378] | [0.003] |
|  | **Low Physical Activity** | **1.197***** | **0.013***** | **4.888***** | **0.051***** |
|  | **(min)** | [0.116] | [0.001] | [0.260] | [0.003] |
|  | **Moderate to Vigorous** | **0.382***** | **0.008***** | **1.345***** | **0.029***** |
|  | **Physical Activity (min)** | [0.056] | [0.001] | [0.124] | [0.003] |
| ****p***<.05; *****p***<.005; ******p***<.0005 | | | | | |

The associations between consecutive days of short sleep and wellbeing indicators were similar to results presented in Table 2, when the Caffeine covariate was removed. The pSS model was preferred for all indicators.

#### Table S3. Personalized vs Fixed Definitions of Short Sleep: LMM Adjusted for Nap Duration

| **Marker ~ 1 + Night in Sequence + Age + Sex + Caffeine + Nap TST + (1\|Subject) + (1\|Subject: Sequence ID)** | | | | | |
| --- | --- | --- | --- | --- | --- |
|  |  | **Fixed Short Sleep (6.0h)** | | **Personalized Short Sleep (P75 TST – 1.0h)** | |
| **Category** | **Marker** | ***B*** [*SE*] | ***β*** [*SE*] | ***B*** [*SE*] | ***β*** [*SE*] |
| ***Mental Wellbeing Markers*** | **Stress** | **0.210***** | **0.009***** | **0.857***** | **0.035***** |
|  | **(Range: 0-100)** | [0.028] | [0.001] | [0.062] | [0.003] |
|  | **Motivation** | **-0.314***** | **-0.012***** | **-0.512***** | **-0.019***** |
|  | **(Range: 0-100)** | [0.025] | [0.001] | [0.055] | [0.002] |
|  | **Sadness** | **0.064**** | **0.004**** | **0.212***** | **0.012***** |
|  | **(Range: 0-100)** | [0.020] | [0.001] | [0.046] | [0.003] |
| ***Sleep Markers*** | **Sleep Satisfaction** | **-0.022***** | **-0.022***** | **-0.117***** | **-0.121***** |
|  | **(Range: 1-5)** | [0.001] | [0.001] | [0.003] | [0.003] |
|  | **Alertness** | **-0.411***** | **-0.016***** | **-0.964***** | **-0.039***** |
|  | **(Range: 0-100)** | [0.022] | [0.001] | [0.048] | [0.002] |
| ***Cardiovascular Markers*** | **Sleep Heartrate** | 0.012 | 0.001 | **0.158***** | **0.019***** |
|  | **(bpm)** | [0.006] | [0.001] | [0.015] | [0.002] |
|  | **Heartrate Variability** | **-0.037*** | **-0.002*** | **-0.284***** | **-0.013***** |
|  | **(rMSSD; ms)** | [0.016] | [0.001] | [0.041] | [0.002] |
| ***Physical Activity Markers*** | **Sedentary** | **1.555***** | **0.012***** | **6.196***** | **0.048***** |
|  | **(min)** | [0.173] | [0.001] | [0.416] | [0.003] |
|  | **Low Physical Activity** | **1.136***** | **0.012***** | **5.694***** | **0.060***** |
|  | **(min)** | [0.119] | [0.001] | [0.290] | [0.003] |
|  | **Moderate to Vigorous** | **0.365***** | **0.008***** | **1.588***** | **0.034***** |
|  | **Physical Activity (min)** | [0.060] | [0.001] | [0.144] | [0.003] |
| ****p***<.05; *****p***<.005; ******p***<.0005 | | | | | |

The associations between consecutive days of short sleep and wellbeing indicators were similar to results presented in Table 2, when nap duration (Nap TST) was added as a covariate. The pSS model was preferred for all indicators.

#### Table S4. Personalized vs Fixed Definitions of Short Sleep: LMM Adjusted for SRI

| **Marker ~ 1 + Night in Sequence + Age + Sex + Caffeine + SRI + (1\|Subject) + (1\|Subject: Sequence ID)** | | | | | |
| --- | --- | --- | --- | --- | --- |
|  |  | **Fixed Short Sleep (6.0h)** | | **Personalized Short Sleep (P75 TST – 1.0h)** | |
| **Category** | **Marker** | ***B*** [*SE*] | ***β*** [*SE*] | ***B*** [*SE*] | ***β*** [*SE*] |
| ***Mental Wellbeing Markers*** | **Stress** | **0.196***** | **0.008***** | **0.795***** | **0.033***** |
|  | **(Range: 0-100)** | [0.028] | [0.001] | [0.062] | [0.003] |
|  | **Motivation** | **-0.334***** | **-0.012***** | **-0.607***** | **-0.022***** |
|  | **(Range: 0-100)** | [0.025] | [0.001] | [0.055] | [0.002] |
|  | **Sadness** | **0.065**** | **0.004**** | **0.223***** | **0.013***** |
|  | **(Range: 0-100)** | [0.020] | [0.001] | [0.046] | [0.003] |
| ***Sleep Markers*** | **Sleep Satisfaction** | **-0.022***** | **-0.023***** | **-0.118***** | **-0.123***** |
|  | **(Range: 1-5)** | [0.001] | [0.001] | [0.003] | [0.003] |
|  | **Alertness** | **-0.428***** | **-0.017***** | **-1.045***** | **-0.042***** |
|  | **(Range: 0-100)** | [0.022] | [0.001] | [0.048] | [0.002] |
| ***Cardiovascular Markers*** | **Sleep Heartrate** | **0.013*** | **0.002*** | **0.165***** | **0.020***** |
|  | **(bpm)** | [0.006] | [0.001] | [0.015] | [0.002] |
|  | **Heartrate Variability** | **-0.037*** | **-0.002*** | **-0.287***** | **-0.013***** |
|  | **(rMSSD; ms)** | [0.016] | [0.001] | [0.040] | [0.002] |
| ***Physical Activity Markers*** | **Sedentary** | **1.221***** | **0.010***** | **4.242***** | **0.033***** |
|  | **(min)** | [0.177] | [0.001] | [0.425] | [0.003] |
|  | **Low Physical Activity** | **0.996***** | **0.010***** | **4.912***** | **0.052***** |
|  | **(min)** | [0.120] | [0.001] | [0.291] | [0.003] |
|  | **Moderate to Vigorous** | **0.322***** | **0.007***** | **1.376***** | **0.030***** |
|  | **Physical Activity (min)** | [0.060] | [0.001] | [0.144] | [0.003] |
| ****p***<.05; *****p***<.005; ******p***<.0005 | | | | | |

The associations between consecutive days of short sleep and wellbeing indicators were similar to results presented in Table 2, when sleep regularity index (SRI) was added as a covariate. The pSS model was preferred for all indicators.

#### Table S5. Personalized vs Fixed Definitions of Short Sleep: P75 – 1.5h vs 5.5h

| **Marker ~ 1 + Night in Sequence + Age + Sex + Caffeine + (1\|Subject) + (1\|Subject: Sequence ID)** | | | | | |
| --- | --- | --- | --- | --- | --- |
|  |  | **Fixed Short Sleep (5.5h)** | | **Personalized Short Sleep (P75 TST – 1.5h)** | |
| **Category** | **Marker** | ***B*** [*SE*] | ***β*** [*SE*] | ***B*** [*SE*] | ***β*** [*SE*] |
| ***Mental Wellbeing Markers*** | **Stress** | **0.360***** | **0.015***** | **1.043***** | **0.043***** |
|  | **(Range: 0-100)** | [0.048] | [0.002] | [0.102] | [0.004] |
|  | **Motivation** | **-0.498***** | **-0.018***** | **-1.221***** | **-0.045***** |
|  | **(Range: 0-100)** | [0.042] | [0.002] | [0.091] | [0.003] |
|  | **Sadness** | **0.128***** | **0.007***** | **0.468***** | **0.027***** |
|  | **(Range: 0-100)** | [0.035] | [0.002] | [0.077] | [0.004] |
| ***Sleep Markers*** | **Sleep Satisfaction** | **-0.033***** | **-0.034***** | **-0.195***** | **-0.203***** |
|  | **(Range: 1-5)** | [0.002] | [0.002] | [0.005] | [0.005] |
|  | **Alertness** | **-0.715***** | **-0.029***** | **-1.780***** | **-0.071***** |
|  | **(Range: 0-100)** | [0.038] | [0.002] | [0.080] | [0.003] |
| ***Cardiovascular Markers*** | **Sleep Heartrate** | **0.025*** | **0.003*** | **0.290***** | **0.035***** |
|  | **(bpm)** | [0.010] | [0.001] | [0.025] | [0.003] |
|  | **Heartrate Variability** | **-0.110***** | **-0.005***** | **-0.543***** | **-0.025***** |
|  | **(rMSSD; ms)** | [0.026] | [0.001] | [0.066] | [0.003] |
| ***Physical Activity Markers*** | **Sedentary** | **1.181***** | **0.009***** | **2.854***** | **0.022***** |
|  | **(min)** | [0.287] | [0.002] | [0.726] | [0.006] |
|  | **Low Physical Activity** | **1.261***** | **0.013***** | **4.842***** | **0.051***** |
|  | **(min)** | [0.195] | [0.002] | [0.485] | [0.005] |
|  | **Moderate to Vigorous** | **0.360***** | **0.008***** | **1.684***** | **0.036***** |
|  | **Physical Activity (min)** | [0.096] | [0.002] | [0.239] | [0.005] |
| ****p***<.05; *****p***<.005; ******p***<.0005 | | | | | |

The associations between consecutive days of short sleep and wellbeing indicators were largely similar to results presented in Table 2, when a stricter TST threshold was used. The pSS model was preferred for all indicators.

#### Table S6. Personalized vs Fixed Definitions of Short Sleep: P80 – 1.0h vs 6.0h

| **Marker ~ 1 + Night in Sequence + Age + Sex + Caffeine + (1\|Subject) + (1\|Subject: Sequence ID)** | | | | | |
| --- | --- | --- | --- | --- | --- |
|  |  | **Fixed Short Sleep (6.0h)** | | **Personalized Short Sleep (P80 TST – 1.0h)** | |
| **Category** | **Marker** | ***B*** [*SE*] | ***β*** [*SE*] | ***B*** [*SE*] | ***β*** [*SE*] |
| ***Mental Wellbeing Markers*** | **Stress** | **0.196***** | **0.008***** | **0.733***** | **0.030***** |
|  | **(Range: 0-100)** | [0.028] | [0.001] | [0.051] | [0.002] |
|  | **Motivation** | **-0.334***** | **-0.012***** | **-0.445***** | **-0.016***** |
|  | **(Range: 0-100)** | [0.025] | [0.001] | [0.045] | [0.002] |
|  | **Sadness** | **0.065**** | **0.004**** | **0.214***** | **0.012***** |
|  | **(Range: 0-100)** | [0.020] | [0.001] | [0.038] | [0.002] |
| ***Sleep Markers*** | **Sleep Satisfaction** | **-0.022***** | **-0.023***** | **-0.092***** | **-0.096***** |
|  | **(Range: 1-5)** | [0.001] | [0.001] | [0.002] | [0.002] |
|  | **Alertness** | **-0.428***** | **-0.017***** | **-0.812***** | **-0.033***** |
|  | **(Range: 0-100)** | [0.022] | [0.001] | [0.039] | [0.002] |
| ***Cardiovascular Markers*** | **Sleep Heartrate** | **0.013*** | **0.002*** | **0.142***** | **0.017***** |
|  | **(bpm)** | [0.006] | [0.001] | [0.012] | [0.001] |
|  | **Heartrate Variability** | **-0.037*** | **-0.002*** | **-0.247***** | **-0.012***** |
|  | **(rMSSD; ms)** | [0.016] | [0.001] | [0.033] | [0.002] |
| ***Physical Activity Markers*** | **Sedentary** | **1.218***** | **0.010***** | **4.773***** | **0.037***** |
|  | **(min)** | [0.177] | [0.001] | [0.344] | [0.003] |
|  | **Low Physical Activity** | **0.999***** | **0.011***** | **4.492***** | **0.047***** |
|  | **(min)** | [0.120] | [0.001] | [0.237] | [0.002] |
|  | **Moderate to Vigorous** | **0.323***** | **0.007***** | **1.195***** | **0.026***** |
|  | **Physical Activity (min)** | [0.060] | [0.001] | [0.118] | [0.003] |
| ****p***<.05; *****p***<.005; ******p***<.0005 | | | | | |

The associations between consecutive days of short sleep and wellbeing indicators were largely similar to results presented in Table 2, when P80 was used instead of P75 for the TST threshold. The pSS model was preferred for all indicators.

#### Table S7. Personalized vs Fixed Definitions of Short Sleep: P80 – 1.5h vs 5.5h

| **Marker ~ 1 + Night in Sequence + Age + Sex + Caffeine + (1\|Subject) + (1\|Subject: Sequence ID)** | | | | | |
| --- | --- | --- | --- | --- | --- |
|  |  | **Fixed Short Sleep (5.5h)** | | **Personalized Short Sleep (P80 TST – 1.5h)** | |
| **Category** | **Marker** | ***B*** [*SE*] | ***β*** [*SE*] | ***B*** [*SE*] | ***β*** [*SE*] |
| ***Mental Wellbeing Markers*** | **Stress** | **0.360***** | **0.015***** | **0.891***** | **0.037***** |
|  | **(Range: 0-100)** | [0.048] | [0.002] | [0.084] | [0.003] |
|  | **Motivation** | **-0.498***** | **-0.018***** | **-0.854***** | **-0.031***** |
|  | **(Range: 0-100)** | [0.042] | [0.002] | [0.074] | [0.003] |
|  | **Sadness** | **0.128***** | **0.007***** | **0.293***** | **0.017***** |
|  | **(Range: 0-100)** | [0.035] | [0.002] | [0.063] | [0.004] |
| ***Sleep Markers*** | **Sleep Satisfaction** | **-0.033***** | **-0.034***** | **-0.155***** | **-0.161***** |
|  | **(Range: 1-5)** | [0.002] | [0.002] | [0.004] | [0.004] |
|  | **Alertness** | **-0.715***** | **-0.029***** | **-1.342***** | **-0.054***** |
|  | **(Range: 0-100)** | [0.038] | [0.002] | [0.065] | [0.003] |
| ***Cardiovascular Markers*** | **Sleep Heartrate** | **0.025*** | **0.003*** | **0.220***** | **0.027***** |
|  | **(bpm)** | [0.010] | [0.001] | [0.020] | [0.002] |
|  | **Heartrate Variability** | **-0.110***** | **-0.005***** | **-0.429***** | **-0.020***** |
|  | **(rMSSD; ms)** | [0.026] | [0.001] | [0.053] | [0.002] |
| ***Physical Activity Markers*** | **Sedentary** | **1.181***** | **0.009***** | **4.706***** | **0.037***** |
|  | **(min)** | [0.287] | [0.002] | [0.583] | [0.005] |
|  | **Low Physical Activity** | **1.261***** | **0.013***** | **5.006***** | **0.053***** |
|  | **(min)** | [0.195] | [0.002] | [0.392] | [0.004] |
|  | **Moderate to Vigorous** | **0.360***** | **0.008***** | **1.707***** | **0.037***** |
|  | **Physical Activity (min)** | [0.096] | [0.002] | [0.193] | [0.004] |
| ****p***<.05; *****p***<.005; ******p***<.0005 | | | | | |

The associations between consecutive days of short sleep and wellbeing indicators were largely similar to results presented in Table 2, when P80 was used instead of P75 for the TST threshold. The pSS model was preferred for all indicators.

#### Table S8. Personalized vs Fixed Definitions of Short Sleep: Associations Between P80 – 1.0h vs 6.0h Proportion of Short Sleep and Cardiometabolic Markers

| **Marker ~ 1 + Proportion of Short Sleep + Age + Sex + Caffeine** | | | | |
| --- | --- | --- | --- | --- |
|  | **Fixed Short Sleep (6.0h)** | | **Personalized Short Sleep (P80 TST – 1.0h)** | |
| **Marker** | ***B*** [*SE*] | ***β*** [*SE*] | ***B*** [*SE*] | ***β*** [*SE*] |
| **Body Roundness Index** | **0.125***** | **0.096***** | **0.179***** | **0.137***** |
|  | [0.028] | [0.021] | [0.047] | [0.036] |
| **Pulse Wave Velocity (m/s)** | 0.017 | 0.013 | **0.105***** | **0.078***** |
|  | [0.025] | [0.019] | [0.042] | [0.031] |
| **Systolic BP  (mmHg)** | 0.548 | 0.039 | **1.521***** | **0.110***** |
|  | [0.291] | [0.021] | [0.481] | [0.035] |
| **Arterial Stiffness Composite** | 0.024 | 0.036 | **0.057***** | **0.085***** |
|  | [0.013] | [0.020] | [0.022] | [0.032] |
| ****p***<.05; *****p***<.005; ******p***<.0005 | | | | |

The associations between proportion of short sleep and cardiometabolic markers were similar to results presented in Table 3 when P80 was used instead of P75 for the TST threshold.

#### Table S9. Personalized vs Fixed Definitions of Short Sleep: Associations Between P75 – 1.5h vs 5.5h Proportion of Short Sleep and Cardiometabolic Markers

| **Marker ~ 1 + Proportion of Short Sleep + Age + Sex + Caffeine** | | | | |
| --- | --- | --- | --- | --- |
|  | **Fixed Short Sleep (5.5h)** | | **Personalized Short Sleep (P75 TST – 1.5h)** | |
| **Marker** | ***B*** [*SE*] | ***β*** [*SE*] | ***B*** [*SE*] | ***β*** [*SE*] |
| **Body Roundness Index** | **0.134***** | **0.103***** | **0.191***** | **0.147***** |
|  | [0.032] | [0.024] | [0.053] | [0.041] |
| **Pulse Wave Velocity (m/s)** | 0.024 | 0.018 | **0.100*** | **0.074*** |
|  | [0.029] | [0.021] | [0.047] | [0.035] |
| **Systolic BP  (mmHg)** | 0.644 | 0.046 | **1.541**** | **0.111**** |
|  | [0.332] | [0.024] | [0.545] | [0.039] |
| **Arterial Stiffness Composite** | **0.030*** | **0.046*** | **0.064*** | **0.096*** |
|  | [0.015] | [0.022] | [0.024] | [0.037] |
| ****p***<.05; *****p***<.005; ******p***<.0005 | | | | |

The associations between proportion of short sleep and cardiometabolic markers were similar to results presented in Table 3 when a stricter TST threshold was used.

#### Table S10. Personalized vs Fixed Definitions of Short Sleep: Associations Between P80 – 1.5h vs 5.5h Proportion of Short Sleep and Cardiometabolic Markers

| **Marker ~ 1 + Proportion of Short Sleep + Age + Sex + Caffeine** | | | | |
| --- | --- | --- | --- | --- |
|  | **Fixed Short Sleep (5.5h)** | | **Personalized Short Sleep (P80 TST – 1.5h)** | |
| **Marker** | ***B*** [*SE*] | ***β*** [*SE*] | ***B*** [*SE*] | ***β*** [*SE*] |
| **Body Roundness Index** | **0.134***** | **0.103***** | **0.160***** | **0.123***** |
|  | [0.032] | [0.024] | [0.046] | [0.035] |
| **Pulse Wave Velocity (m/s)** | 0.024 | 0.018 | **0.101*** | **0.075*** |
|  | [0.029] | [0.021] | [0.041] | [0.030] |
| **Systolic BP  (mmHg)** | 0.644 | 0.046 | **1.517**** | **0.109**** |
|  | [0.332] | [0.024] | [0.468] | [0.034] |
| **Arterial Stiffness Composite** | **0.030*** | **0.046*** | **0.061**** | **0.091**** |
|  | [0.015] | [0.022] | [0.021] | [0.031] |
| ****p***<.05; *****p***<.005; ******p***<.0005 | | | | |

The associations between proportion of short sleep and cardiometabolic markers were similar to results presented in Table 3 when a stricter TST threshold was used.

#### Table S11. Number and Demographic details of Cross-Country Sample

| Country | Number | Age (median & IQR) | % Female |
| --- | --- | --- | --- |
| Australia | 9859 | 42 (18) | 58.28 |
| Finland | 37696 | 46 (19) | 55.18 |
| Germany | 16212 | 43 (18) | 56.72 |
| Japan | 6532 | 43 (16) | 28.58 |
| Singapore | 1968 | 42 (13) | 49.92 |
| UK | 10636 | 43 (18) | 57.21 |
| USA | 293855 | 40 (21) | 71.45 |

#### Table S12. Country Level Sleep Characteristics

| Country | Bedtime | TST | Weekday TST | Weekend TST | PSS Proportion | Weekday PSS Proportion | Weekend PSS Proportion |
| --- | --- | --- | --- | --- | --- | --- | --- |
| Australia | 22:29 [103.98] | 7.17 [1.41] | 7.11 [1.36] | 7.34 [1.53] | 29.75 [15.04] | 30.64 [17.27] | 25 [17.82] |
| Finland | 23:11 [101.12] | 7.13 [1.4] | 7.03 [1.32] | 7.44 [1.53] | 29.53 [14.82] | 31.84 [17.65] | 21.25 [16.26] |
| Germany | 23:08 [104.48] | 7.05 [1.43] | 6.94 [1.37] | 7.38 [1.56] | 30.79 [14.81] | 33.04 [18.24] | 21.92 [16.15] |
| Japan | 23:54 [137.57] | 6.38 [1.65] | 6.29 [1.58] | 6.61 [1.8] | 36.51 [17.23] | 38.1 [20.04] | 30 [19.09] |
| Singapore | 23:52 [121.03] | 6.72 [1.53] | 6.64 [1.47] | 6.95 [1.67] | 33.13 [16.08] | 34.44 [19.06] | 26.92 [18.46] |
| UK | 22:59 [96.97] | 7.13 [1.37] | 7.06 [1.31] | 7.37 [1.48] | 28.31 [15.04] | 29.73 [17.89] | 22.38 [16.72] |
| USA | 22:48 [111.03] | 7.16 [1.51] | 7.08 [1.46] | 7.38 [1.63] | 31.95 [15.82] | 33.33 [18.35] | 25.81 [17.98] |

### Values shown are Median [IQR];

### The Singaporean sample here does not overlap with our research sample, is slightly older but obtains more sleep than our sample. The average sleep depicted here is longer than for a sample of working adults from a population health study; 6.3h; bedtime 00:15 (Ong JL, Sleep (2021) zsaa179: <https://doi.org/10.1093/sleep/zsaa179>)

### Supplementary Methods

#### Filtering criteria for main sleep

A single main sleep (sleep period with longest TST) was identified between 18:00 the previous day to 17:59 on the current day. To accurately characterize the diversity of sleep patterns under free-living conditions – including irregular or short sleep and nights without sleep – data loss due to non-wear or technical faults were accounted for as follows.

The main sleep episode may be missed if extended periods of non-wear occurred during habitual sleep hours, leading to underestimation of TST{Ahmadi, 2020 #132}. Habitual sleep patterns were quantified using the empirical mean sleep propensity at each 15min time window *t* of the day, defined as the proportion of days on which sleep was detected within that time window:

$\hat{p}\left( t \right)=\frac{1}{D}\sum_{d=1}^{D} S_{d}(t)$ , where $S_{d}\left( t \right)=\left\{ \begin{aligned} 1, \text{asleep at time }\text{t}\text{ on day }\text{d} \\ 0, &\text{otherwise} \end{aligned} \right.$

$S_{d}(t)$ included any sleep detected, i.e., not restricted to main sleep. Using sleep propensity rather than fixed clock time accommodates individual differences in sleep timing and irregular sleep schedules. A night was excluded if, weighted by habitual sleep propensity, the expected sleep during non-wear exceeded that of sleep detected during observed wear:

$\sum_{t=1}^{T} \hat{p}(t)N_{d}(t)>\sum_{t=1}^{T} \hat{p}(t)S_{d}(t)$ , where $N_{d}\left( t \right)=\left\{ \begin{aligned} 1, \text{non-wear at time }\text{t}\text{ on day }\text{d} \\ 0, &otherwise \end{aligned} \right.$

Nights were not excluded because sleep was short, absent, or occurred at non-habitual times per se. Rather, exclusion was applied when non-wear during likely sleep periods created uncertainty regarding whether the main sleep episode had been missed. Nights with long non-wear periods, particularly during habitual sleep hours, were therefore excluded unless a sleep episode compatible with habitual sleep was detected. Conversely, if the Oura Ring was worn during habitual sleep hours, a night would generally be retained even if sleep was absent, short, fragmented, or occurred during non-habitual hours. This approach permitted non-habitual sleep patterns while minimizing false classification of short sleep resulting from missed main sleep periods. 7,144 (5.25%) nights were excluded as a result.

SS runs with missing baseline values were excluded from analyses as the run history could not be determined. After excluding 5 [7] (median [IQR]) fSS runs and 5 [6] pSS runs preceded by missing nights, there were 48 [41] fSS and 55 [30] pSS runs remaining. Of these remaining runs, the night after 4 [6] fSS runs and 4 [5] pSS runs were missing and excluded from recovery night analyses.

Any other sleep episode with at least 10min TST were considered naps. Total nap duration between 18:00 the previous day to 17:59 on the current day were used in analyses.

#### Cross-Country Data Collection

The multi-country supporting data to establish the generalizability of the proposed framework was provided by Ouraring Inc. (San Francisco, CA) according to their terms and conditions ([https://ouraring.com/terms-and-conditions](https://www.google.com/url?q=https://ouraring.com/terms-and-conditions&sa=D&source=docs&ust=1779796473340786&usg=AOvVaw0ri1df2rHVosJJfA4z0t3G)), and privacy policy ([https://ouraring.com/en/privacy-policy-oura-health](https://www.google.com/url?q=https://ouraring.com/en/privacy-policy-oura-health&sa=D&source=docs&ust=1779796473340889&usg=AOvVaw0SFNmF6rBktFlhBvkr6NDJ)). Data used for this study were anonymized or aggregated consistent with Oura’s privacy policy. The data come from users in 7 countries with different sleep habits (Australia, Finland, Germany, Japan, Singapore, the United Kingdom and the United States of America). Data from 430,386 users were collected from 1st March 2024 to 28th February 2025. Mean age was 42.6 years (SD=13.2), mean BMI was 25.8 (SD=4.9) and the proportion of females was 66.2% (see Table S11). Users were required to have at least 100 nights of valid sleep recordings to be included. Details about sleep habits are presented in Table S12.
